# Germline pathogenic variation impacts somatic alterations and patient outcomes in pediatric CNS tumors

**DOI:** 10.1101/2025.02.04.25321499

**Authors:** Ryan J. Corbett, Rebecca S. Kaufman, Shelly W. McQuaid, Zalman Vaksman, Saksham Phul, Miguel A. Brown, Jennifer L. Mason, Sebastian M. Waszak, Bo Zhang, Chuwei Zhong, Emily Blauel, Heena Desai, Ryan Hausler, Ammar S. Naqvi, Jessica M. Daggett, Alex Sickler, Evan C. Cresswell-Clay, Patricia J. Sullivan, Antonia Chroni, Zhuangzhuang Geng, Elizabeth M. Gonzalez, Yuankun Zhu, Allison P. Heath, Marilyn Li, Penn Medicine BioBank, Regeneron Genetics Center, Phillip B. Storm, Adam C. Resnick, Kara N. Maxwell, Kristina A. Cole, Angela J. Waanders, Miriam Bornhorst, Suzanne P. MacFarland, Jo Lynne Rokita, Sharon J. Diskin

## Abstract

The contribution of rare pathogenic/likely pathogenic (P/LP) germline variants to pediatric central nervous system (CNS) tumor development remains understudied. Here, we characterized the prevalence and clinical significance of germline P/LP variants in cancer predisposition genes across 830 CNS tumor patients from the Pediatric Brain Tumor Atlas (PBTA). We identified germline P/LP variants in 23.3% (193/830) of patients and the majority (137/193) lacked clinical reporting of genetic tumor syndromes. Among P/LP carriers, 34.6% had putative somatic second hits or loss of function tumor alterations. Finally, we linked pathogenic germline variation with novel somatic events and patient survival to highlight the impact of germline variation on tumorigenesis and patient outcomes.

## Introduction

Central nervous system (CNS) tumors are the leading cause of childhood cancer death in the United States, with over 47,000 children and young adults diagnosed annually worldwide^1,2^. The overall prevalence of children with cancer with reported rare pathogenic or likely pathogenic (P/LP) germline variants in cancer predisposition genes (CPGs) ranges from 4-27%^3–14^, although comprehensive studies in pediatric cohorts with CNS tumor malignancies are limited. There are over 20 characterized genetic cancer predisposition syndromes (CPS) observed in patients with CNS tumors (**Table S1**), and loss-of-function variants in over 30 CPGs have been linked to increased risk of distinct pediatric CNS tumor histologies and molecular subtypes^15^. Targeted germline sequencing is being increasingly implemented in clinical settings to inform clinical practice and treatment strategies^16–18^. However, the full spectrum and prevalence of pathogenic germline variants in CPGs in patients with CNS tumors remains unknown, limiting the clinical utility of targeted testing.

The integration of germline and somatic multi-omic modalities with clinical data in cancer patients has the potential to uncover novel mechanisms underlying tumor biology and reveal clinical outcomes driven by genetic predisposition^19–21^. For example, rare pathogenic germline variants in CPGs in patients with medulloblastoma have been linked to molecular subgroups, specific evolutionary trajectories, and/or biological pathway deficiencies, including chromothripsis (associated with *TP53* variants), homology-directed repair deficiency (*PALB2* and *BRCA2*), and proteome instability (*ELP1*)^22,23^.

In this study, we investigate rare pathogenic germline variants in CPGs across 830 pediatric CNS tumor patients in the Pediatric Brain Tumor Atlas (PBTA), consisting of individuals enrolled on either the Children’s Brain Tumor Network (CBTN) or a Pediatric Neuro-Oncology Consortium (PNOC) protocol^24^. We integrate multiple data modalities from matched tumors^25,26^, including DNA and RNA sequencing, proteomics, DNA methylation, and clinical data. We observe higher prevalence of CPG germline P/LP variants than previously reported in PBTA patients, with enrichment in PBTA relative to two cancer-free control cohorts. Importantly, we identify CPS-associated pathogenic germline variants in patients with undiagnosed tumor predisposition syndromes, underscoring the need for implementing germline testing at diagnosis. Finally, we report new associations between rare germline pathogenic CPG variants and somatic alterations in patients with high-grade glioma, *BRAF* fusion-positive low-grade gliomas, and medulloblastoma.

## Results

### Cohort characteristics and study workflow

For this study, we analyzed germline sequencing data from 830 pediatric CNS tumor patients with matched tumor sequencing, including 790 with whole genome sequencing (WGS) and 40 with whole exome sequencing (WXS) (**Figure 1A, Table S2**). Tumors were classified into 15 broad histology groups using the 2021 World Health Organization CNS tumor classifications^27^, and molecular subtypes were assigned using annotations from the Open Pediatric Cancer Project^26^ (**Figure 1B**). We further distinguished diffuse intrinsic pontine gliomas (DIPG) and diffuse midline gliomas (DMG) from other high-grade gliomas (HGG) due to their distinct anatomical, molecular, and clinical profiles. Demographic and genomic characteristics, including genetically-inferred sex and ancestry from germline data, and tumor-specific alterations, are summarized in **Figures 1C** and **S1**. Rare germline variants (allele frequency <0.1% in all non-bottleneck populations in gnomAD non-cancer population database^28^) were evaluated for pathogenicity, and we focused specifically on variants in 211 cancer predisposition genes (CPGs) defined by an expert panel of physicians and scientists (**Table S3**). Germline P/LP variants were integrated with matched tumor data, including RNA-Seq (N = 770, 92.8%), proteomics (N=192; 23.1%), and DNA methylation array data (N=730; 88.0%). Sixty-seven patients (8.1%) had documented CPS, with the most prevalent being neurofibromatosis type 1 (NF-1; N=25), followed by tuberous sclerosis (N=10), neurofibromatosis type 2 (NF-2; N=7), and Li-Fraumeni syndrome (N=7) (**Table S2**).

**Figure 1.**
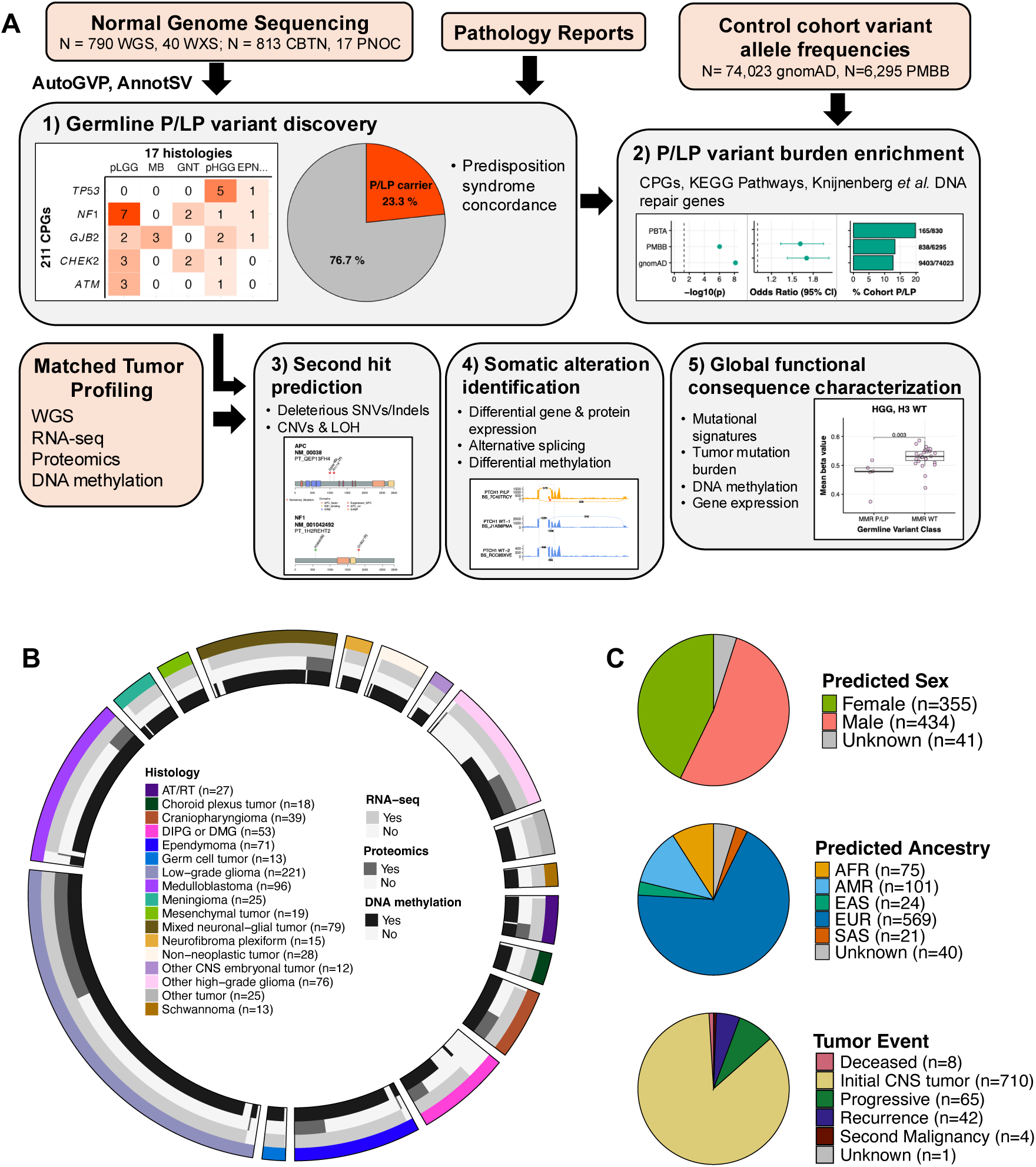
Pediatric CNS tumor cohort germline analysis summary. **A.** Overview of germline and integrative somatic analyses. **B.** Circos plot summarizing distribution of tumor histologies and availability of matched tumor RNA-seq, proteomics, and DNA methylation data modalities in 830 pediatric CNS tumor patients. Atypical Teratoid Rhabdoid Tumors (AT/RT, N=27); choroid plexus tumors (CPT, N=18); craniopharyngiomas (N=39); diffuse intrinsic pontine glioma or diffuse midline glioma (DIPG or DMG, N=53); ependymoma (EPN, N=71); germ cell tumors (GCT, N=13); low-grade gliomas (LGG, N=221); medulloblastomas (MB, N=96); meningiomas (MNG, N=25); mesenchymal tumors (N=19); mixed glioneuronal and neuronal tumors (GNT, N=79); neurofibroma plexiforms (NFP, N=15); other non-AT/RT, non-MB CNS embryonal tumors (N=12); other high-grade gliomas (HGG, N=76); and schwannomas (SWN, N=13). All other benign tumors and non-cancerous lesions were assigned to a “non-neoplastic tumor” category (N=28), and other rare CNS tumors of low sample size (N<10) were grouped into an “Other tumor” category (N=25). **C.** Sex predicted genetic ancestry superpopulation, and matched tumor event distributions among cohort. AFR=sub-Saharan African, AMR=admixed American, EAS=East Asian, EUR=European, SAS=South Asian.

### Rare P/LP germline variants in CPGs are prevalent in pediatric CNS tumor patients

Using AutoGVP^29^, we identified 197 rare P/LP germline SNVs/InDels in 71 CPGs (**Figure 2A**, **Figure S2A**, **Table S4A-B**), with 80.2% supported by ClinVar evidence (N=158/197; **Figure 2B**). We also detected 18 germline CPG structural variants (SVs) classified as P/LP by AnnotSV^30^ or ClassifyCNV^31^ (**Figure 2C**, **Table S4C**). To validate these findings and further characterize their clinical relevance, we cross-referenced CPG P/LP variants with known CNS CPS (**Table S1**). We identified 104 variants in 100 patients, including 48 variants in 48 distinct patients (5.7% of cohort) without a clinically-reported CPS (**Figure 2D**, **Table S5**). These findings may reflect previously unrecognized germline contributions to tumor development and highlight the potential value of systematic germline analysis in uncovering clinically-meaningful variation beyond known diagnoses. Among patients with a clinically reported CPS in the Open Pediatric Cancer (OpenPedCan) Project^26^, 86% (49/57) had a matching P/LP germline variant in our study (**Figure S2B**). In the remaining eight cases, we extended our search to include low allele frequency variants (0.08-0.20 VAF) and variants of uncertain significance (VUS), uncovering potentially deleterious variants in five patients (**Table S6**). These included four VUS: one *TP53* missense variant in an LFS patient (PT_PFP1ZVHD), and one *NF1* and two *NF2* splice donor variants in patients with neurofibromatosis. We also recovered a P/LP *NF2* variant with a VAF of 0.175, leading us to identify nine additional low-VAF P/LP variants (**Table S7**).

**Figure 2.**
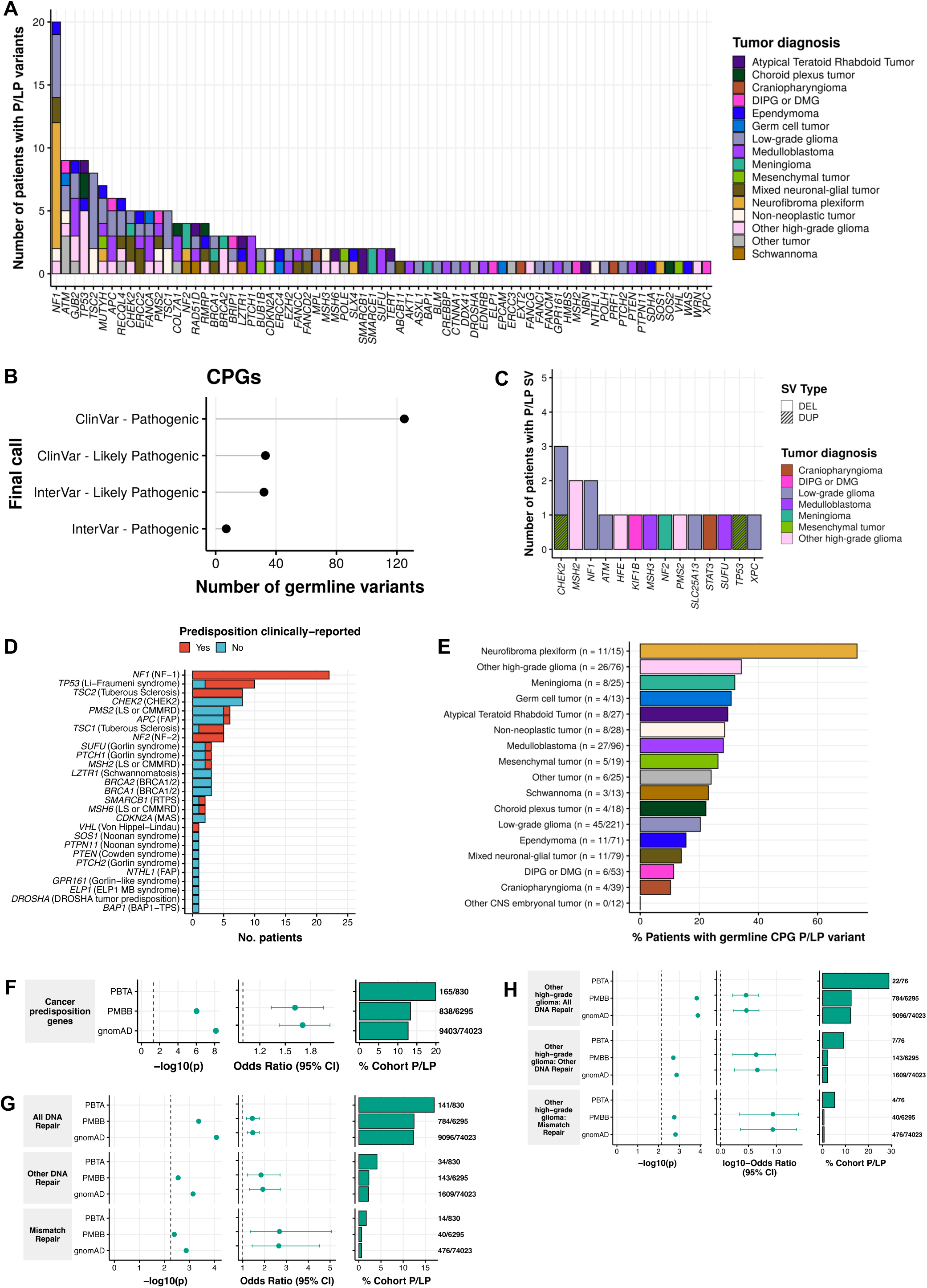
Prevalence of cancer predisposition gene germline P/LP variants in pediatric CNS tumor patients. **A.** Number of patients with identified germline P/LP variants by cancer predisposition gene (CPG) and tumor histology. **B.** Number of CPG germline P/LP SNVs and InDels identified by call and call source. **C.** Number of patients with identified P/LP structural variants by gene, tumor histology, and type. DEL=deletion, DUP=duplication. . Number of patients harboring P/LP variants in syndrome-associated genes, and clinical definition. RTPS=Rhabdoid Tumor Predisposition Syndrome, FAP=Familial Adenomatous Polyposis, MAS=melanoma astrocytoma syndrome, BAP1 TPS=BAP1 Tumor Predisposition Syndrome. **E.** Percent of patients harboring a CPG P/LP variant by tumor histology. **F.** Odds ratios and associated p-values of CPG P/LP variant burden among pediatric CNS tumor cohort relative to PMBB and gnomAD cancer-free control cohorts. **G-H.** Odds ratios and associated p-values of P/LP variant burden in DNA repair genes from Knijnenburg *et al.* 2018 among **G)** entire cohort and **H)** pediatric high-grade glioma cohort (excluding DIPG and DMG) relative to control cohorts. Dashed lines in p-value plots indicate Bonferroni-adjusted p<0.05.

Overall, P/LP CPG variants were identified in 23.3% (193/830) of patients, including 30 with two variants and two with three variants: one patient with HGG and three *MSH6* P/LP variants and another with subependymal giant cell astrocytoma (SEGA, PT_JW6FBEFK) that harbored *TSC2*, *ASXL1*, and *APC* P/LP variants (**Table S4A**). The former was clinically diagnosed with constitutional mismatch repair deficiency syndrome (CMMRD; PT_3CHB9PK5), suggesting biallelic *MSH6* activation (**Figure S2C**). In the latter SEGA case, the *ASXL1* variant is unlikely to represent clonal hematopoiesis of indeterminate potential (CHIP): the patient was four years old, the variant had higher VAF in tumor vs. blood (0.60 vs. 0.41), and no other CHIP-associated variants were found. No homozygous P/LP variants were observed (**Figure S2D**); however, in addition to the CMMRD patient with compound heterozygous *MSH6* variants, a patient with medulloblastoma (MB, PT_2FK75B27) had two adjacent *PMS2* P/LP InDels on the same allele (**Figure S2E**), despite no reported CPS.

P/LP carriers were non-randomly distributed across histologies (p=5.0e-4, **Table 1),** and enriched in neurofibroma plexiform (NF; n=11/15, OR=9.5, 95% CI=2.7-41.0, p=4.6e-06) and HGG (n=26/76, OR=1.8, 95% CI=1.1-3.2, p=0.02) cohorts (**Figure 2E**, **Figure S3A**). Enrichment was also observed among tumor molecular subtypes with known genetic predispositions (p<0.05, OR lower 95% CI>1.0; **Figure S3B, Table S8**). All SEGA patients (N=10) had P/LP variants, primarily in *TSC1* (n=3) or *TSC2* (n=6). The remaining SEGA patient had a *MUTYH* P/LP variant, not previously linked to SEGA. SHH-activated MB (SHH-MB) patients were enriched for P/LP carriers (n=12/19), including variants in *PTCH1* (n=3), *SUFU* (n=3), and *GPR161* (n=1). Among histone H3 wildtype HGGs, 64% (n=16/25) of patients were P/LP carriers, mainly in mismatch repair (MMR) genes (n=6) or *TP53* (n=4). Pineoblastoma (PB) patients were significantly enriched for P/LP variants (n=4/5; **Figure S3C**). None harbored *DICER1* variants known to be associated with PB^32^, but one had a *DROSHA* and two had *ATM* P/LP variants. Demographic features were not associated with P/LP carrier status (**Table 1, Table S9**), except among mixed glioneuronal and neuronal tumors (GNTs), in which male carriers were overrepresented (n=10 male vs. 1 female, OR=9.8, 95% CI=1.3-445.5, p=0.02; **Table S9C**). In total, we identified 225 germline CPG P/LP variants (n=207 SNVs/InDels, n=18 SVs) in 23.3% of pediatric CNS tumor patients.

**Table 1.** Demographic and clinical features of PBTA cohort by CPG P/LP carrier status.

|  | **CPG P/LP** | **No CPG P/LP** | **p** |
| --- | --- | --- | --- |
| **Number Cohort** | **193 (23.3%)** | **637 (76.8%)** |  |
| **Sex** |  |  | **0.21** |
| Female | 93 (48.2%) | 262 (45.1%) |  |
| Male | 96 (49.7%) | 338 (53.4%) |  |
| Unknown | 4 (2.1%) | 37 (5.8%) |  |
| **Race** |  |  | **0.34** |
| American Indian or Alaska Native | 0 (0%) | 5 (0.8%) |  |
| Asian | 2 (1%) | 25 (3.9%) |  |
| Black or African American | 14 (7.3%) | 53 (8.3%) |  |
| More Than One Race | 2 (1%) | 9 (1.4%) |  |
| Native Hawaiian or Other Pacific Islander | 0 (0%) | 1 (0.2%) |  |
| Other/Unknown Race | 36 (18.7%) | 99 (15.5%) |  |
| White | 139 (72.0%) | 445 (69.9%) |  |
| **Ethnicity** |  |  | **0.74** |
| Hispanic or Latino | 22 (11.4%) | 62 (9.7%) |  |
| Not Hispanic or Latino | 163 (84.5%) | 544 (85.4%) |  |
| Other/Unknown Ethnicity | 8 (4.1%) | 31 (4.9%) |  |
| **Genetic Ancestry Superpopulation** |  |  | **0.84** |
| AFR | 18 (9.3%) | 57 (8.9%) |  |
| AMR | 28 (14.5%) | 73 (11.5%) |  |
| EAS | 4 (2.1%) | 20 (3.1%) |  |
| EUR | 134 (69.4%) | 435 (68.3%) |  |
| SAS | 5 (2.6%) | 16 (2.5%) |  |
| Unknown | 4 (2.1%) | 36 (5.7%) |  |
| **Tumor Histology** |  |  | **5.0E-04** |
| ATRT | 8 (4.1%) | 19 (3.0%) |  |
| Choroid plexus tumor | 4 (2.1%) | 14 (2.2%) |  |
| Craniopharyngioma | 4 (2.1%) | 35 (5.5%) |  |
| DIPG or DMG | 8 (4.1%) | 45 (7.1%) |  |
| Ependymoma | 11 (5.7%) | 60 (9.4%) |  |
| Germ cell tumor | 4 (2.1%) | 9 (1.4%) |  |
| Low-grade glioma | 47 (24.4%) | 174 (27.3%) |  |
| Medulloblastoma | 28 (14.5%) | 68 (10.7%) |  |
| Meningioma | 8 (4.1%) | 17 (2.7%) |  |
| Mesenchymal tumor | 5 (2.6%) | 14 (2.2%) |  |
| Mixed neuronal-glial tumor | 11 (5.7%) | 68 (10.7%) |  |
| Neurofibroma plexiform | 11 (5.7%) | 4 (0.6%) |  |
| Non-neoplastic tumor | 8 (4.1%) | 20 (3.1%) |  |
| Other CNS embryonal tumor | 0 (0%) | 12 (1.9%) |  |
| Other high-grade glioma | 26 (13.5%) | 50 (7.8%) |  |
| Other tumor | 6 (3.1%) | 19 (3.0%) |  |
| Schwannoma | 4 (2.1%) | 9 (1.4%) |  |
| **Median age at diagnosis, years** | 7.93 | 8.50 | 0.47 |
| **Median tumor mutation burden** | 0.39 | 0.36 | 0.12 |

### Germline pathogenic variation in CPGs and cancer pathways is enriched in pediatric CNS tumor patients

We compared the prevalence of P/LP germline variants in CPGs between pediatric CNS tumor patients and non-cancer cohorts from the Penn Medicine BioBank (PMBB; n=6,295) and gnomAD (n=74,023) (see **Methods**). CPG P/LP variants were significantly enriched relative to PMBB (OR=1.6, 95% CI=1.3-1.9, p=9.4e-07) and gnomAD (OR=1.7, 95% CI=1.4-2.0, p=7.3e-9) (**Figure 2F, Table S10**). Histology-specific analyses revealed significant CPG P/LP variant burden in patients with NF, HGG, and MB compared to controls (Bonferroni-adjusted p<0.05, OR lower 95% CI>1.0; **Figure S4**). Genes with the strongest burden included *NF1*, *TSC2*, *TSC1*, *TP53*, and *NF2* (**Figure S5A**), largely reflecting the composition of the cohort, which includes many patients with LGGs, NF, and HGG. We confirmed known associations, such as *PTCH1* with MB, *SMARCB1* with ATRT, and *SMARCE1* with meningioma (MNG, **Figure S5B**). Notably, we also identified a novel enrichment of *ATM* P/LP variants in patients with PB, though the small sample warrants cautious interpretation (n=2/5 patients).

Expanding beyond CPGs, we assessed rare P/LP variants across all genes and found 13 enriched KEGG pathways in the PBTA, including five involving the previously implicated CPGs (*TP53*, *TSC1*/*2*, and *NF1*). Additional enriched pathways included cell cycle regulation (G1/S) and broader cancer signaling networks such as the *MTOR* pathway (**Figure S6, Table S10C**). We further assessed DNA repair pathways^33^ and observed significant enrichment of P/LP variants across 1) all DNA repair genes, 2) MMR genes, and 3) other DNA repair genes in PBTA (OR=1.4-2.7, 95% CI=1.2-5.0, p<1.3e-03; **Figure 2G, Table S10D**). This is likely driven by patients with HGG with high P/LP burden in DNA repair and MMR genes (OR=2.9-8.7, 95% CI=1.7-25.0, p<2.0e-03; **Figure 2H**), consistent with previous studiess^25,34^. In summary, germline P/LP variants in CPGs are enriched in the PBTA CNS tumor cohort, with a novel potential association between *ATM* P/LP variants and PB.

### Germline CPG P/LP carriers exhibit loss of function (LOF) somatic events

We assessed all matched tumors for evidence of somatic LOF, including SNVs/InDels, copy number (CN) alterations, and loss of heterozygosity (LOH; **Figure 3A, Table S11**). Of the 225 germline P/LP variants, 195/207 (94.2%) SNVs/InDels and 17/18 (94.4%) SVs were detected in tumors. The 12 undetected SNVs/InDels had significantly lower germline VAFs compared to those detected in tumors (median VAF 0.20 vs. 0.50, p=2.9e-07), suggesting potential genetic mosaicism. Among detected variants, 72 events in 70 patients had a putative somatic second hit. These included eight patients with somatic SNVs/InDels annotated as oncogenic or likely oncogenic (O/LO) by OncoKB^35^ or otherwise predicted as deleterious (see **Methods**): *NF1* mutations in four distinct P/LP carriers, an *APC* nonsense mutation and *SUFU* splice site mutation in two MB patients (PT_QEP13FH4 and PT_WWZ2QQ14R, respectively), a *MSH6* missense mutation in the HGG CMMRD patient (PT_3CHB9PK5), and a *SMARCE1* splice site mutation in a MNG case (PT_NSXP8AWV; **Figure 3B, Figure S7**). Next, we identified 63 LOH events (28.0% of all variants), with 55 (87.3%) resulting in loss of the wildtype allele (**Figure 3C**). Sixteen of these were accompanied by CN loss or deep deletions, while the remaining 47 events occurred in CN-neutral tumors (**Figure S8A**). Recurrent LOH was observed in known predisposition contexts, including: *NF1* (LGG, GNT, NF), *NF2* (MNG, SWN), *PTCH1* and *SUFU* (MB), *MSH2* (HGG), *SMARCB1* (ATRT), *TP53* (CPT, HGG), and *TSC1* (LGG)^22,36–38^.

**Figure 3.**
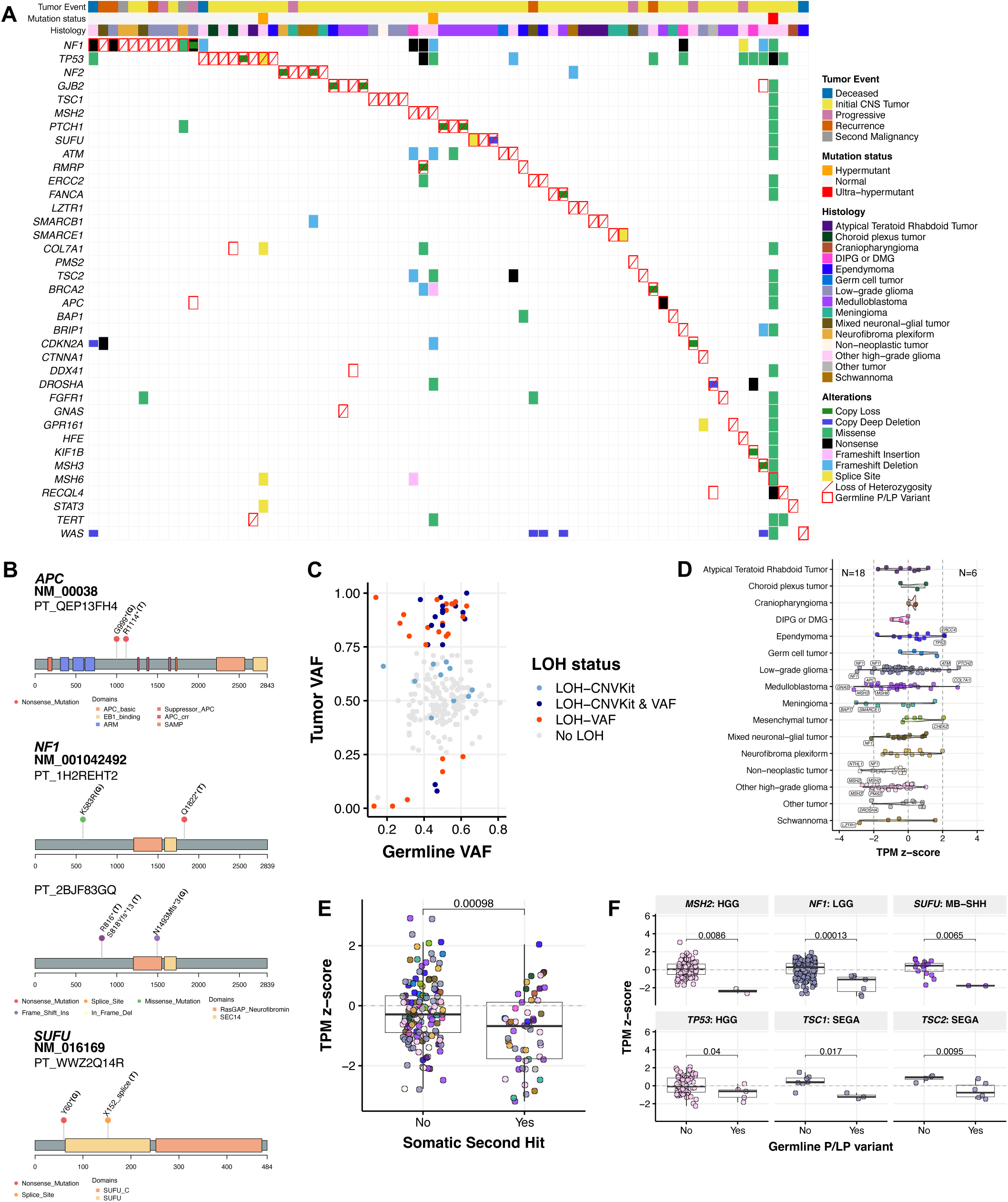
Identification of somatic second hit events in matched tumors. **A.** Oncoprint of somatic alterations (SNVs/InDels, copy number variation, loss of heterozygosity, differential gene and protein expression) in matched tumors of patients harboring CPG P/LP variants. **B.** Lollipop plots displaying germline P/LP variants (“G”) and oncogenic SNVs/InDels (“T”) in matched tumors of *APC*, *NF1*, and *SUFU* P/LP carriers. **C.** Tumor versus germline variant allele frequency (VAF) of all P/LP variants detected in matched tumors. **D.** Violin plot of cancer predisposition gene (CPG) transcripts per million (TPM) z-scores in matched tumors of CPG P/LP carriers by histology. Vertical dashed lines indicate z-score thresholds for significantly increased and decreased gene expression. N=18 and N=6 indicate number of cases exhibiting significant transcript loss and gain, respectively. Dot colors correspond to tumor histology-specific color assignments defined in Figure 1B. **E.** TPM z-scores of CPGs in P/LP carriers with identified somatic second hits versus those with no second hits. P-value is derived from a Wilcoxon rank sum test. **F.** TPM z-scores by histology and CPG in P/LP carriers versus non-carriers, with Wilcoxon rank sum test p-values.

To assess the potential functional impact of germline P/LP variants on gene expression, we surveyed transcriptional and proteomic profiles in matched tumors. We observed 18 cases of gene expression loss and six cases of gene expression gain (**Figure 3D**). Variants associated with gene expression loss were primarily stop-gained (n=7), frameshift (n=3), SV deletions (n=3), or splice variants (n=2) (**Table S11**). In contrast, only two variants associated with expression gain were frameshift, while the remainder were either SV duplications (n=1) or missense variants (n=3), the latter of which are unlikely to directly inhibit transcription. Importantly, carriers with a somatic second hit (i.e., LOF SNV/InDel or wildtype LOH) exhibited significantly reduced transcript abundance relative to P/LP carriers without a second hit (p=9.7e-04; **Figure 3E**). Recurrent transcript loss in P/LP carriers versus non-carriers was observed for *MSH2*, and *TP53* (HGG), *NF1* (LGG), *SUFU* (SHH-MB), and *TSC1* and *TSC2* (SEGA) (Wilcoxon p<0.05, **Figure 3F**). Proteomics analysis of matched tumors revealed protein loss in three P/LP carriers: *TSC2* (PT_66XN3MT1, SEGA) with transcript loss (z-score=-2.43), *SLC25A13* (PT_MPRBGGEJ, LGG), and *ELP1* (PT_2JDDX6TJ, H3K27M DMG) (**Table S11**). While *ELP1* is a known SHH-MB predisposition gene^23^, its role in H3K27M DMG has not been reported. In total, we identified somatic genomic or expression LOF events associated with germline P/LP variation in 78/225 (34.6%) cases.

We also analyzed tumor DNA methylation data to investigate potential epigenetic changes driven by germline P/LP variation. We found 102 hypermethylated and 317 hypomethylated probes annotated to the same gene in matched tumors of P/LP carriers relative to non-carriers (**Figure S8B, Table S12**). P/LP-associated hyper- and hypo-methylation were enriched in gene promoters (OR=1.9, 95% CI=1.2-2.9, p=5.4e-03) and intronic regions (OR=1.5, 95% CI=1.1-1.8, p=3.6E-06), respectively. Promoter hypermethylation was associated with gene expression loss (**Figure S8C**), implicating epigenetic silencing as a secondary mechanism. For example, *RECQL4* promoter hypermethylation was observed in in two unrelated P/LP carriers (LGG and GNT; **Figure S8D-E**), both with marginal gene expression loss (z-scores=-0.89, −1.18) but no somatic mutations. Notably, 14 P/LP carriers exhibited somatic promoter hypermethylation in the absence of other somatic hits or transcript loss, supporting epigenetic inactivation as an additional tumor mechanism driven by germline predisposition.

### P/LP variation is significantly associated with differential somatic alternative splicing and impacts gene expression and protein function

Due to the large number of germline splice variants in the PBTA cohort (37/217) and their documented impact on pediatric CNS tumor development^22,23,39^, we investigated their influence on somatic alternative splicing (AS) and subsequent functional consequences. We queried splice events (see **Methods**) in matched tumors and observed that germline P/LP splice region variation was associated with significantly increased proximal intron retention or exon skipping relative to other P/LP variants (**Figure 4A, Table S13**). Splice region variants were associated with reduced transcript and protein abundance (**Figure S9A-B**), similar to levels observed for frameshift and stop gained variants thus suggesting similar functional effects. Frameshift and stop gained P/LP variants were associated with proximal intron retention and alternative splice site usage, respectively, each significantly negatively correlated with transcript abundance (Pearson’s R=-0.64, p=0.013 and R=-0.38, p=0.036), implicating aberrant splicing as a mechanism influencing gene expression in P/LP carriers (**Figure 4B**).

**Figure 4.**
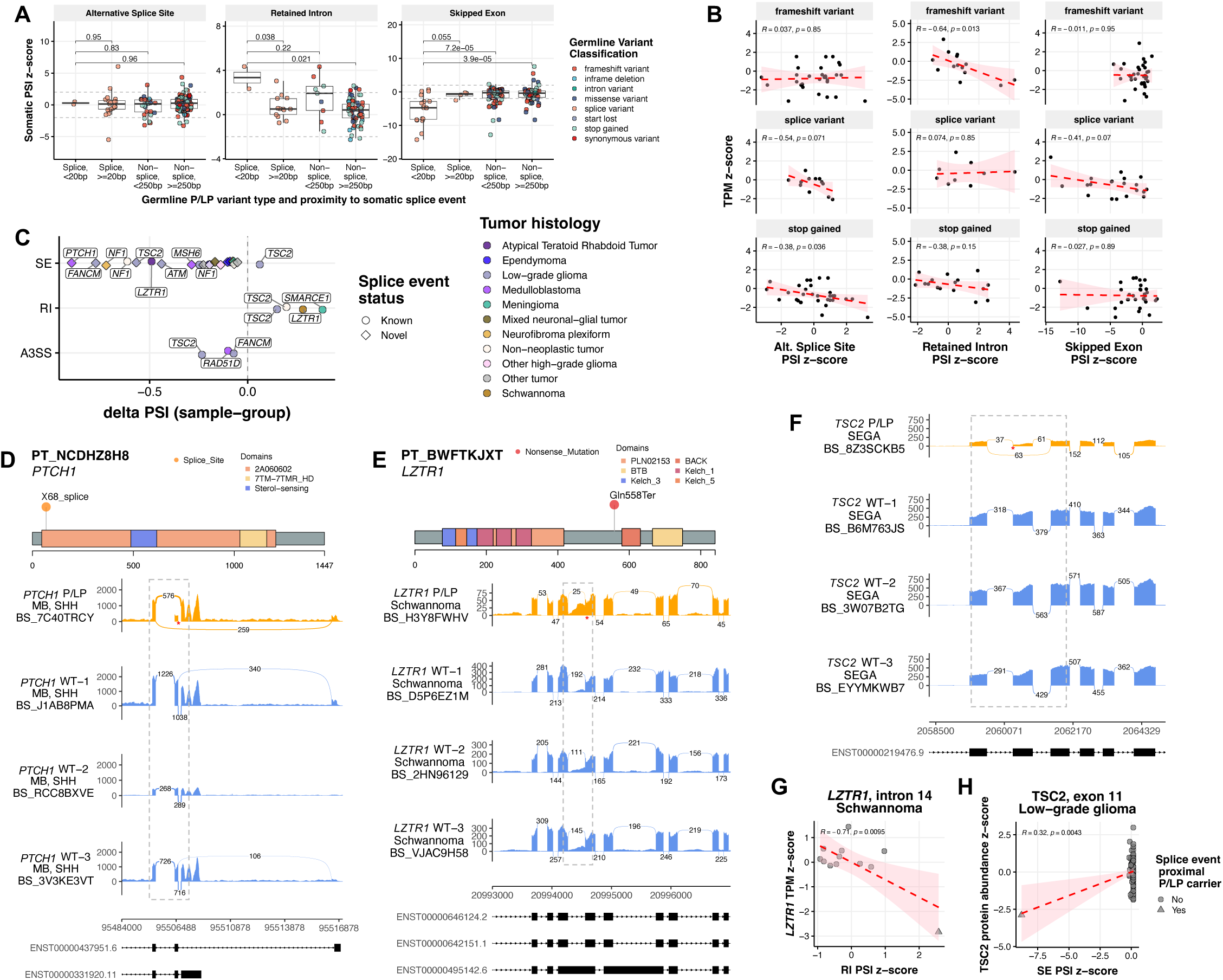
Landscape of germline P/LP variant-associated alternative splicing events in matched tumors. **A.** Germline P/LP variant-proximal splice event percent splice index (PSI) z-scores in matched tumors, grouped by proximity to germline variant and class of variant (splice vs. non-splice). P-values are derived from Wilcoxon rank-sum tests **B.** Transcripts per million (TPM) z-scores against P/LP-variant-proximal splice event PSI z-scores among frameshift variant, splice variant, and stop gained variant P/LP carriers. R values represent Pearson correlation coefficients. **C.** Significant P/LP variant-proximal alternative splicing events in matched tumors of P/LP carriers by splice event type and histology, plotted by percent spliced in (PSI) difference relative to other tumors of same histology. SE=single exon, RI=retained intron, A3SS=alternative 3’ splice site. **D.** A germline *PTCH1* splice acceptor P/LP carrier with SHH-activated medulloblastoma (SHH-MB) exhibits increased exon 2 skipping relative in matched tumor relative to other SHH-MB tumors. **E.** A *LZTR1* stop gained P/LP carrier with schwannoma exhibits increased upstream intron 14 retention in matched tumor relative to other schwannomas. **F.** A *TSC2* splice polypyrimidine tract P/LP variant carrier with subependymal giant cell astrocytoma (SEGA) exhibits increased exon 11 skipping in matched tumor relative to other SEGAs. **G.** *LZTR1* TPM z-scores against intron 14 PSI z-scores in schwannoma cases. **H.** *TSC2* TPM z-scores against exon 11 PSI z-scores in low-grade gliomas. **I.** Two *GBA* splice donor P/LP carriers with *KIAA1549::BRAF* fusion positive LGG and SHH-MB exhibit increased exon 2 skipping in matched tumors relative to other tumors of the same histology. **J.** Indexed tumor mutation burdens in *KIAA1549::BRAF* fusions-positive LGG with *GBA* P/LP carrier PT_EQ5C5TEA highlighted. **K.** *GBA* sashimi plot highlighting exon 2 skipping in initial and metastatic second malignancy of PT_EQ5C5TEA.

We identified 32 significant P/LP variant-proximal AS events in matched tumors (|percent spliced in [PSI] z-score| ≥ 2) including skipped exon (SE; N=25), retained intron (RI; N=4), and alternative 3’ splice site usage (A3SS; N=3) events (**Figure 4C**). Intriguingly, 20 (80%) of the 24 SE events lack annotated splice junctions are therefore considered novel. Several tumors showed AS-associated gene expression loss or putative disruption of Pfam protein domains^40,41^ (**Figure 4D-I, Figure S9C**). In a germline *PTCH1* splice acceptor P/LP carrier with SHH-MB (PT_NCDHZ8H8) we observed novel exon 2 skipping, which affects a conserved sterol transporter domain (**Figure 4D**). A germline *LZTR1* stop gained P/LP carrier with schwannoma (PT_BWFTKJXT) exhibited increased intron 14 retention directly upstream of this variant that was associated with significant transcript loss relative to other schwannomas (R=-0.71, p=9.5e-3; **Figure 4E,G**), suggesting a mechanism of *LZTR1* inactivation. A germline *TSC2* polypyrimidine tract P/LP carrier with SEGA exhibited exon 11 skipping resulting in a novel transcript isoform and significant TSC2 protein loss compared to other LGGs (**Figure 4F,H**). Lastly, a patient with AT/RT and a germline *LZTR1* synonymous variant exhibited somatic exon 4 skipping, predicted to disrupt the Kelch substrate recognition domain (**Figure S9D**). This variant has previously been shown to disrupt an exonic *LZTR1* splice enhancer^42^. We observe significant LOH in this patient’s tumor, implicating a novel role for *LZTR1* splice variants in AT/RT tumorigenesis. These findings support a broad role of germline splice and non-splice variants in shaping the AS landscape and its downstream functional consequences in pediatric CNS tumors.

### Mismatch repair gene pathogenic variation drives distinct genetic and molecular signatures in pediatric high-grade gliomas

Given the significant P/LP variant burden in DNA repair pathways, we further explored their functional impact in tumors. We identified 105 P/LP variants across 31 DNA repair CPGs in 97 patients (11.7% of cohort; **Figure S10A**). HGG tumors were enriched in patients with P/LP variants in MMR, all repair, and other repair pathways (FDR<0.05 and OR lower CI>1 for all comparisons), while mesenchymal and other tumors were nominally enriched among base and nucleotide excision repair (BER and NER) P/LP carriers, respectively (p<0.05 and OR lower CI>1 for all comparisons; **Figure 5A**). We assessed COSMIC mutational signature data from these tumors and confirmed that H3-wildtype HGGs from HR and MMR gene P/LP carriers exhibited significantly higher SBS3 and MMR-deficiency signature exposures, respectively, relative to other HGGs (p=0.029 and 0.0033, **Figure 5B-C**, **Figure S10B, Table S14**). MMR P/LP carriers also harbored significantly higher tumor mutation burden (TMB) compared to non-carriers (p=1.2e-04), and, consistent with previous findings in MMR-deficient HGG^43^, 4/6 tumors were considered hypermutant (>10 mutations/Mb; **Figure 5D**). Mesenchymal tumors with BER P/LP variants exhibited increased SBS30 exposure weights relative to non-carriers (p=8.6e-4, **Figure 5E).** We observed significantly lower genome-wide mean methylation beta values in H3-wildtype HGGs from MMR P/LP carriers compared to non-carriers, with the CMMRD patient with 3 *MSH6* germline P/LP variants exhibiting the lowest global methylation (p=0.003, **Figure 5F**). Since the tumors of MMR P/LP carriers were hypomethylated and hypermutated, we asked whether TMB and global methylation were broadly correlated in HGGs. Indeed, global methylation was significantly negatively correlated with TMB exclusively in H3 wildtype HGG tumors (Pearson’s R=-0.72, p=7.6e-06), but not in other histologies or HGG subtypes (**Figure S10C-D, Table S15**). Clustering of HGG tumors by beta values at the 20,000 most variable promoter CpGs revealed a distinct cluster of heterozygous MMR P/LP carriers, while the CMMRD patient sample clustered separately (**Figure 5G**). Finally, two MMR gene P/LP carriers exhibited somatic promoter hypermethylation of the same gene, suggestive of epigenetic silencing associated with P/LP variation despite observed global hypomethylation (**Figure 5H-I**). These findings highlight distinct DNA methylation and mutational signatures associated with germline MMR P/LP variation in pediatric HGGs.

**Figure 5.**
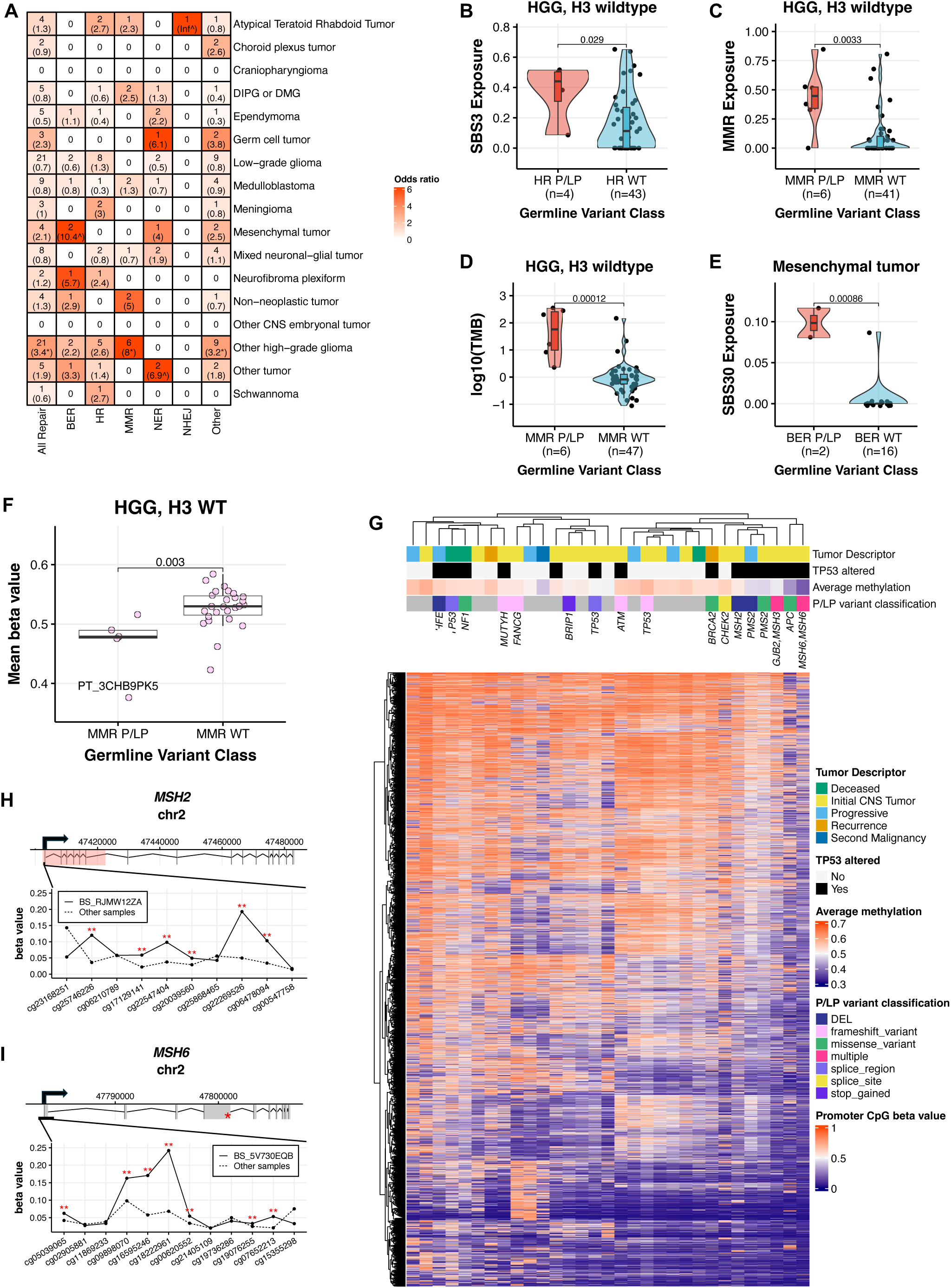
Germline P/LP variants in DNA repair genes are associated with distinct molecular profiles in high-grade glioma (HGG). **A.** Count of DNA repair variant P/LP carriers within CNS tumor histology cohorts by DNA repair pathway, with Fisher’s exact test-derived odds ratios in parentheses. Cells are colored by odds ratio weight. *=FDR<0.05, ^=p<0.05. BER=base excision repair, HR=homologous recombination, MMR=mismatch repair, NER=nucleotide excision repair, NHEJ=non-homologous end joining. “Other” indicates genes in the DNA repair list that are not in any of the five pathways. **B.** SBS3 mutational signature exposure weights in HR gene P/LP carriers versus non-carriers with H3-wildtype HGG. **C-D.** MMR deficiency mutational signatures exposure weights **(C)** and tumor mutation burden **(D)** in MMR gene P/LP carriers versus non-carriers with H3-wildtype HGG. **E.** SBS30 exposure weights in BER gene P/LP carriers versus non-carriers with mesenchymal tumors. **F.** Mean beta value from all probes on Infinium MethylationEPIC array in MMR gene P/LP carriers versus non-carriers with H3-wildtype HGG, with constitutional mismatch repair deficiency (CMMRD) syndrome patient indicated by label. P-values are derived from Wilcoxon rank sum tests. **G.** Clustering of H3 wildtype HGG samples by 20k most variable promoter-annotated probes on the Infinium MethylationEPIC array. Gene symbols above heatmap indicate annotation of germline P/LP variants, and *MSH6/MSH6* indicates homozygous P/LP carrier. Hierarchical clustering was performed using the Euclidean distance measure and ward D2 clustering method. **H-I.** A germline P/LP deletion in *MSH2* **(H)** and P/LP SNP in *MSH6* **(I)** are associated with significant hypermethylation of respective promoter probes in H3-wildtype HGG cases. The red box and asterisk in gene models indicate positions of germline P/LP deletion and SNP, respectively. **=|beta value z-score| ≥ 2.

### CPG P/LP carriers exhibit distinct survival outcomes

CPG P/LP variants have previously been linked to poorer overall survival (OS) in pediatric cancers including neuroblastoma and leukemia^22,44–46^. We therefore evaluated whether P/LP carriers in the PBTA exhibited distinct survival outcomes (**Figure 6A, Table S16**). We found that among all *KIAA1549*::*BRAF* fusion-positive LGGs, P/LP carriers had significantly worse event-free survival (EFS) versus non-carriers in univariate (p=0.031, **Figure 6B**) and multivariate cox proportional hazards models accounting for extent of tumor resection, predicted sex, and age at diagnosis (HR=2.67, p=0.03; **Figure 6C**). Most P/LP carriers with *KIAA1549::BRAF* fusion positive LGG (N=13/20) harbored variants in DNA repair genes, and this cohort exhibited the second greatest excess DNA repair P/LP variant burden relative to controls; this represent a novel finding, but was not significant after multiple test correction (OR=1.8, 95% CI=1.1-3.0, p=0.01; **Figure S11A**).

**Figure 6.**
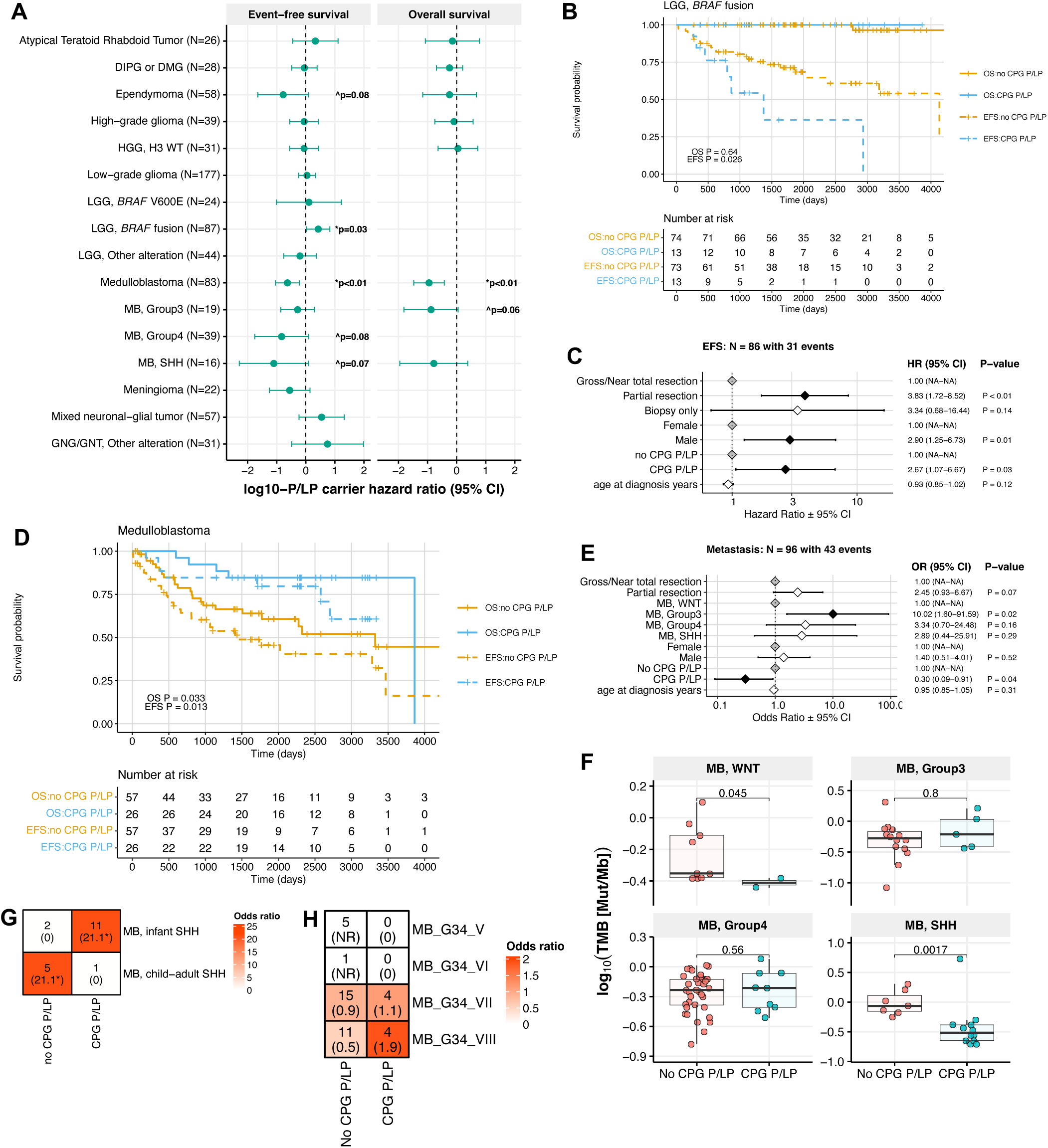
P/LP carrier status-associated survival differences in pediatric CNS tumor patients. **A.** Hazard ratios of event-free and overall survival (EFS and OS, respectively) in germline CPG P/LP carriers versus non-carriers by tumor histology or molecular subtype. Statistics were derived from cox proportional hazards models that included age at diagnosis covariate for all tumor histologies, and molecular subtype and extent of tumor resection covariates for relevant tumor histologies. **B.** Kaplan-Meier survival curves for EFS and OS in patients with *KIAA1549::BRAF* fusion-positive LGG, stratified by CPG P/LP carrier status. **C.** Cox proportional hazards model forest plot of EFS in patients with *KIAA1549::BRAF* fusion positive LGG, including covariates for extent of surgical resection, age at diagnosis, and interaction between pathology diagnosis and CPG P/LP carrier status. **D.** Kaplan-Meier survival curves for EFS and OS in patients with medulloblastoma, stratified by CPG P/LP carrier status. **E.** Logistic regression model forest plot of metastasis in patients with medulloblastoma. **F.** Tumor mutation burden in patients with medulloblastoma by CPG P/LP carrier status and molecular subtype. P-values are derived from Wilcoxon rank sum tests. **G-H.** Heatmap of patient distribution among **H)** SHH-MB subgroups and **I)** Group 4 MB methylation subgroups by CPG P/LP carrier status, with Fisher’s exact test-derived odds ratios in parentheses. *=FDR<0.05. NR=not reportable.

Among patients with MB, CPG P/LP carriers exhibited significantly better EFS and OS relative to non-carriers (p<0.01 for both EFS and OS in univariate and multivariate models; **Figure 6D, Figure S11B-C**). We evaluated additional molecular and clinical data in patients with MB, and observed that P/LP carriers exhibited significantly lower metastasis rates compared to non-carriers (OR=0.31, 95% CI=0.09-0.91, p=0.04; **Figure 6E**), and metastasis was associated with significantly worse OS (HR=7.88, 95% CI=2.1-29.1, p=4.2e-03; **Figure S11D**). We identified significantly lower TMB in WNT- and SHH-MB tumors of P/LP carriers compared to non-carriers, which has not been previously reported (**Figure 6F**). In the case of SHH-MB tumors, this may be explained in part by earlier ages at diagnosis in P/LP carriers (**Figure S11E**). We further delineated SHH-MB tumors and observed that infant SHH (β and γ) tumors were enriched in P/LP carriers (N=6, OR=1.7, 95% CI=1.4-1395.7, p=0.01; **Figure 6G**), while child-adult SHH (α and δ) tumors were enriched in non-carriers and exhibited worse OS and EFS **(Figure S11F)**. Similarly, we only identified P/LP carriers with subgroup 7 (N=3/19) and 8 (N=3/15) Group 4 MBs among this subtype, and these patients had significantly better EFS and OS than non-carriers with subgroup 5 tumors, though we acknowledge the low numbers in these groups (**Figure 6H, Figure S11G**). In summary, P/LP carriers with MB Group 4 subgroups 7 or 8, infant-type SHH MB, and/or MB with decreased metastasis confer better OS.

## Discussion

We identified germline CPG P/LP variants in 193/830 (23.3%) patients, a frequency that is within the range reported in previous germline studies (4-27%)^3–5,7,8,11–13,22^. While the majority of CPG P/LP SNVs/InDels called by AutoGVP utilized ClinVar evidence, we identified 42 additional P/LP variants through AutoGVP’s modified InterVar classification, 18 P/LP SVs and one deep intronic *NF1* variant previously classified as P/LP^47^, highlighting the importance of leveraging multiple approaches to characterize the full spectrum of disease-causing germline variation in pediatric cancers. We observed high concordance between clinically-reported CPSs and prevalence of associated CPG P/LP variants, and identified several VUS in patients lacking P/LP variants that warrant further study into their pathogenicity. The majority of identified germline P/LP variants were absent from clinical records, including a high proportion of variants in genes linked to CPSs. Several factors may contribute to this, such as the extended timeframe of sample collection (spanning over 13 years), the non-routine nature of germline testing, and a negative family history due to incomplete penetrance or *de novo* variants^48^. These findings underscore the importance of increasing routine germline testing in pediatric CNS tumor patients to better guide monitoring and treatment strategies even in the absence of family history. For example, we identified several patients with germline P/LP *TP53* or MMR gene variants with no clinically reported CPS; the former would benefit from lifetime monitoring for early detection of secondary cancers as is routine for patients with LFS, and the latter could receive immunotherapies given the prevalence of chemoresistant tumors^49^. While the majority of histology-specific P/LP CPGs have been previously described, we observed that 2 of 5 patients with PB harbored an *ATM* germline P/LP variant. To our knowledge, *ATM* has not been implicated in development of PB, although future studies are needed to assess their prevalence in a larger cohort.

We detected a putative somatic second hit (LOF SNV/InDel, LOH) in tumors of 27.5% (N=53/193) of P/LP carriers, suggesting that observed pathogenic germline variation in these patients is indeed contributing to tumorigenesis. While representing roughly one-third of cases, and within the range of second hit frequencies reported in previous pediatric cancer studies (10-55%)^3–5,8,44^, there may be additional somatic inactivating mutations in functionally-related genes and/or pathways, and/or haploinsufficiency alone may initiate tumorigenesis^50^ in some cases. Furthermore, a small percentage of germline CPG P/LP variants (13/225) were not detected in matched tumors, and these exhibited low median VAF (0.2) relative to those variants found in tumors (VAF=0.5). Such observations may suggest that these low VAF variants represent genetic mosaicism and may not contribute to tumorigenesis; however, it is possible that these variants play critical roles in tumor initiation and be lost at later stages of tumor evolution, as has been reported in glioma and medulloblastoma initiating events^51,52^. Somatic SNVs/InDels in the same P/LP gene were rare, while CN-neutral LOH resulting in loss of wildtype allele was pervasive, particularly in *NF1*, *TP53*, *PTCH1*, *MSH2*, and *TSC2*.

We report transcriptomic, regulatory, and proteomic somatic alterations indicative of gene inactivation, many in cases in which a tumor DNA second hit was not observed. The majority of somatic differential gene expression in CPG P/LP carriers indicated loss of transcript in the same gene, though there were rare cases of gene expression gain including in a *CHEK2* P/LP carrier with a large germline duplication event. For the remaining cases, increased gene expression associated with P/LP germline variation may be due in part to compensatory upregulation as a result of loss of function at the protein level; however, we did not have proteomics data from any of these samples to confirm protein loss. We further identified splice region P/LP variants driving somatic aberrant splicing and disruption of protein domains with subsequent gene expression and protein loss. Beyond splice region variants, we report cases of frameshift or stop gained variant-associated AS, indicating that splicing may contribute to LOF driven by these variant classifications as has been reported previously^53^. Our work emphasizes the need for further investigation into the contributions of AS to pediatric CNS tumor development and progression. Lastly, we observed global functional consequences of germline MMR gene P/LP variation in patients with histone H3-wildtype HGG, including increased MMR-deficiency mutational signatures, TMB, and global hypomethylation, the latter of which has not been reported previously. Strikingly, we found increased TMB to be associated with global hypomethylation exclusively in H3-wildtype HGG, but not in other HGG subtypes nor in other tumor histologies.

Germline CPG pathogenic variation was associated with distinct clinical outcomes in a subset of patients. CPG P/LP carriers with *KIAA1549::BRAF* fusion-positive LGGs exhibited worse EFS compared to non-carriers. Interestingly, this cohort disproportionately harbored DNA repair gene variants, which were also nominally enriched among *KIAA1549::BRAF*-fusion positive LGGs relative to control populations. Future research should seek to assess the contribution of DNA repair gene germline pathogenic variation to *KIAA1549::BRAF* LGG tumorigenesis. Conversely, CPG P/LP carriers with MB exhibited better survival relative to non-carriers, which may be explained by lower metastasis rates and TMB, earlier ages at diagnosis, and clinically favorable methylation subgroups associated with P/LP carriers in this MB cohort. While inactivation of known MB predisposition genes has previously been associated with increased metastasis risk^54,55^, this has not been explored in the context of germline variation to our knowledge. Future studies with a larger cohort are required to assess unique germline variant-associated tumor characteristics that may contribute to clinical outcomes in MB. In summary, our study has identified new functional links between germline variation, tumor molecular features, and clinical outcomes in pediatric CNS tumor patients, highlighting the importance of routine germline testing at diagnosis. Continued understanding of germline susceptibility and its influence on tumorigenesis will aid in identification of patients that may benefit from genetic counseling, surveillance, altered treatment regimens, and ultimately clinical outcomes.

## Methods

### Pediatric CNS tumor/normal samples and germline SNV/indel calling

The pediatric brain tumor cohort used in this study is composed of patients from the Children’s Brain Tumor Network (CBTN, https://cbtn.org/, n=775) and the Pediatric Neuro-Oncology Consortium (PNOC, https://pnoc.us/, n=55). The CBTN is a collaborative international and multi-institutional research and biorepository initiative dedicated to the study of childhood brain tumors^24^. PNOC is an international clinical trials consortium dedicated to bringing new therapies to children and young adults with brain tumors. Together, these consortia have contributed samples to the Childhood Cancer Data Initiative (dbGaP phs002517.v4.p2). Germline variant calls from whole genome sequencing (WGS) data for paired tumor (∼60X) and normal peripheral blood (∼30X)^25,26^ and from whole exome sequencing (WXS) data for paired tumor (∼470X) and normal peripheral blood (∼308X) were obtained through an approved dbGaP data access request. Matched tumor WGS (∼60X), WXS (∼470X), RNA-Seq, methylation, and/or proteomics somatic outputs as well as clinical and demographic information (e.g. survival, metastasis, race, ethnicity, and clinically-reported predisposition syndrome) from the OpenPedCan^26^ v15 data release^56^ were utilized in this study. The list of genes associated with central nervous system (CNS) tumor predisposition syndromes was compiled using two key sources: Chapter 14 of the 2021 World Health Organization (WHO) Classification of Tumors of the Central Nervous System^57^ and the recent publication describing *DROSHA*-associated tumor predisposition^58^. These references collectively informed the gene set used to identify P/LP germline variants relevant to CNS tumor syndromes in our cohort. We also performed pathology and electronic health record review (CHOP IRB 09-007316) for all patients with P/LP variants in CNS tumor predisposition associated CPGs to identify whether a tumor syndrome was clinically reported.

### Germline SNV/indel calling

Briefly, paired-end WGS and WXS reads were aligned to the version 38 patch release 12 of the Homo sapiens reference genome using BWA-MEM^59^. The Broad Institute’s Best Practices^60^ were used to process Binary Alignment/Map files (BAMs) in preparation for variant discovery. Duplicate alignments were marked using SAMBLASTER^61^, and merged and sorted BAMs using Sambamba^62^. The BaseRecalibrator submodule of the Broad’s Genome Analysis Tool Kit (GATK)^63^ the GATKHaplotypeCaller^64^ submodule to generate a genomic variant call format (GVCF) file. This file was used as the basis for germline calling. Germline haplotype calling was performed using the GATK Joint Genotyping Workflow on individual gVCFs from the germline sample alignment workflow. The GATK generic hard filter suggestion was applied to the VCF (SNPs only), with an additional requirement of 10 reads minimum depth per variant. This Kids First workflow can be found at https://github.com/kids-first/kf-germline-workflow.

### Cancer predisposition gene list curation

Cancer predisposition genes (CPGs) were selected based on review of the Children’s Hospital of Philadelphia’s (CHOP) Division of Genomic Diagnostics’ hereditary cancer panel (www.testmenu.com/chop/Tests/786446), public databases including the Online Mendelian Inheritance in Man (www.omim.org) and the National Institute of Health’s Genetics Home Reference (www.medlineplus.gov/genetics), and medical literature. We added *GJB2* due to its observed enrichment in neuroblastoma compared to tumor-free control cohorts^65^ (**Table S3**).

### Germline variant annotation and assessment of variant pathogenicity

Resulting variants were annotated using the Kids First Data Resource Center (KFDRC) Germline SNV Annotation Workflow and added the following annotations: ENSEMBL 105, ClinVar (2022May07), and InterVar (https://github.com/kids-first/kf-germline-workflow/blob/master/docs/GERMLINE_SNV_ANNOT_README.md). Germline variants in 211 cancer predisposition genes were further analyzed.

Pathogenicity was assessed for filtered variants *in silico* using AutoGVP to evaluate ClinVar (2022May07) and a modified execution of InterVar^29^. We applied a pathogenicity pre-processing workflow on VEP-annotated VCFs that encompasses variant annotation with ANNOVAR (database pulled on 2024May06), InterVar v2.2.1 variant classification, and AutoPVS1 v2.0.0 variant scoring. Variants with read depth coverage ≥ 10, variant allele fraction ≥ 0.20, and observed in <0.1% across non-bottleneck gnomAD v3.1.1 non-cancer populations (African/African American, Admixed American, non-Finnish European, South Asian, East Asian) were retained for variant classification. Pathogenicity assessment was performed using AutoGVP as previously described^29^. Briefly, we pulled classifications for rare variants annotated in the ClinVar database with ≥2 stars or with 1 star and no conflicting submissions. All variants with conflicting classifications in ClinVar were resolved by first filtering for submissions that have been associated with MedGen disease concept IDs, and taking the single submission or consensus submission if available. Remaining variants classifications were resolved by taking the classification from the submission at the last date evaluated. For remaining variants without ClinVar annotation, we adjusted PP5 based on this modified InterVar assessment and corrected PVS1 using AutoPVS1. Variants were assigned as pathogenic (P), likely pathogenic (LP), benign (B), likely benign (LB), or unknown significance (VUS) by first considering ClinVar results and then the modified InterVar output. This approach was applied to pediatric CNS cases and non-cancer control samples (PMBB and gnomAD 3.1.1). For variants with conflicting submissions in ClinVar that were resolved as non-P/LP, we reviewed cases for which there was a P or LP submission that provided functional evidence (i.e., experimental evidence showing this variant is associated with loss of function), and upgraded the classification accordingly when such evidence was sufficient.

All cases of proximal adjacent InDels in a single patient and InDels ≥ 20 bp were manually reviewed in Integrated Genomics Viewer (IGV) and removed if variants were not present. We also queried samples for intronic NF1 variants identified in Koczkowska et al. 2023^47^ that were shown to associate with alternative splicing events and classified these as LP.

### Cancer-free control cohorts

We utilized cancer-free control cohort data from the Penn Medicine BioBank (PMBB) and Genome Aggregation Database (gnomAD v3.1) to calculate relative enrichment of P/LP variants in the PBTA cohort. The PMBB is a precision medicine cohort that began in 2004, enrolling patients from the University of Pennsylvania Health System who consented to biospecimen collection and linkage of their samples to electronic health record (EHR) data. Whole-exome sequencing of participant DNA extracted from stored buffy coats was performed by the Regeneron Genetics Center (Tarrytown, NY). For this study, we leveraged data from 6,295 cancer-and tumor-free individuals classified based on the absence of relevant International Classification of Diseases (ICD)-9/10 codes in their EHRs, as previously described^65^.

### Germline structural variant calling and pathogenicity assessment

We applied the KFDRC Germline Structural Variant Caller Workflow to generate SV pathogenicity calls from normal sequencing data (https://github.com/kids-first/kf-germline-workflow/blob/master/docs/GERMLINE_SV_README.md). Briefly, Manta v1.6.0^66^ was run on all aligned paired-end sequencing reads to call SVs, and AnnotSV v3.1.1^30^ was run on Manta output to annotate SV pathogenicity. CNVnator^67^ was run on aligned reads to call CNVs (https://github.com/kids-first/kf-germline-workflow/blob/master/docs/GERMLINE_CNV_README.md), and ClassifyCNV^31^ was run on CNVnator output to annotate CNV pathogenicity. We filtered out any P/LP SVs or CNVs present in >1% of the PBTA cohort, and any that overlapped with SV or CNV annotated in gnomAD at an allele frequency > 0.01% in a non-bottleneck population.

### Cohort demographics and clinical characteristics

Genetic sex prediction was performed as previously described^25^. Briefly, we calculated the fraction of total normalized X and Y chromosome reads that were attributed to the Y chromosome. Values < 0.2 were annotated as female, values > 0.4 were predicted male, and samples with values between 0.2–0.4, inclusive, were marked as unknown. Genetic ancestry prediction was performed on all patients with normal WGS data (n=790) as previously described^68^. Differences in demographic and clinical characteristics between subjects with and without germline P/LP variants in CPGs were assessed across the cohort and within cancer histologies. Fisher’s exact tests were used to compare categorical data variables (sex, race, ethnicity, predicted genetic ancestry) and Mann Whitney-U tests were used to compare numerical data variables (age at diagnosis, overall and event-free survival, tumor mutation burden). A two-sided Fisher’s exact test or Mann Whitney-U test p-value <0.05 was considered significant.

### CPG P/LP Enrichment

The gnomAD v3.1.1 and PMBB cancer-free control cohorts were run through the same pathogenicity preprocessing and assessment pipelines. The PMBB cohort consisted of tumor-free patients without a family history of cancer, as defined previously^44^. Variants identified in the PMBB cohort were subjected to the same manual curation used for the PBTA cohort. Enrichment of P/LP germline variants across all CPGs and within individual CPGs in the PBTA cohort vs. control cohorts was performed using Fisher’s exact tests. The same statistical framework was applied to assess enrichment of P/LP germline variants among genes in KEGG pathways and DNA repair pathways as defined in Knijnenburg et al 2018^33^, regardless of whether the gene was also defined as a CPG. Bonferroni adjustment was performed for gene and pathway analyses to correct for multiple tests. Enrichment of CPGs and pathway P/LP variants was also run within tumor histology cohorts using the same methodology.

### Somatic second hit analyses

Somatic SNVs, CNVs, and SVs were obtained from OpenPedCan release v15^25,26^. Somatic mutations in CPGs were annotated with oncoKB to identify those classified as oncogenic or likely oncogenic (O/LO)^35^. We defined somatic SNV/indel second hits as those defined as those annotated as O/LO or otherwise classified as “probably_damaging” or “possibly_damaging” by PolyPhen or “deleterious” by SIFT. Copy number variants (CNVs) were identified in tumor samples as described in OpenPedCan^25^ based on WXS CNVkit^69^ calls or WGS consensus calls among Control-FREEC^70^, CNVkit^69^, GATK^63^ and/or MantaSV^66^. We assessed LOH in matched tumors using two methods: 1) comparison of VAFs in tumor vs. germline, and 2) estimating tumor allele copy number using CNVkit^69^, the latter of which accounts for low tumor purity which limited detection of LOH by VAF in some cases. We created the publicly-available tool AlleleCouNT (https://github.com/d3b-center/AlleleCouNT)^71^ to obtain germline and tumor allele counts of each CPG P/LP variant in P/LP carriers, and calculated odds ratios of variant frequencies from 2×2 contingency tables using Fisher’s exact tests. Loss of heterozygosity (LOH) affecting the wildtype allele was defined by one of the following criteria: 1) tumor VAF > 0.75, odds ratio > 1 and Fisher’s exact p < 0.05, or 2) CNVkit copy number of wild type allele=0. LOH affecting the variant allele was defined by the following criteria: 1) tumor VAF < 0.25, odds ratio < 1 and Fisher’s exact p < 0.05, or 2) CNVkit copy number of variant allele=0. Variant LOH score was calculated as tumor VAF - germline VAF. We calculated gene-level LOH score using the following equation across all rare variants showing evidence of heterozygosity in the germline:

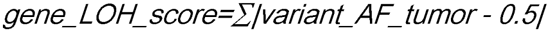

Genes were defined as exhibiting significant LOH if gene_LOH_score > 0.25.

### Gene Expression

RNA-seq data were obtained from the OpenPedCan release v15^26,72^. Z-scores were calculated using the formula *z=(x - μ)=*σ where x is the sample TPM for a given gene, *μ* is the mean gene TPM across samples, and σ is the TPM standard deviation. We considered a gene as differentially expressed relative to other samples of the same tumor histology when |TPM z-score| ≥ 2.

### Alternative Splicing

We queried replicate Multivariate Analysis of Transcript splicing (rMATs) files from OpenPedCan release v15^26,72^ for the most proximal splice event of each case type (skipped exon [SE], retained intron [RI], and alternative 3’ and 5’ splice site [A3SS and A5SS, respectively] to each CPG P/LP variant, and calculated percent spliced in (PSI) value z-scores for these events in each P/LP carrier using the same method we applied to TPM values. Since rMATs only reports splice events in a sample that are either supported by 1) the provided gene annotation file or 2) sample RNA-seq reads, we classified each splice event as known (i.e., annotated in a transcript) or novel (i.e., not found in any annotated transcript). In the case of novel SE and RI events that were not reported in a given sample, we set PSI equal to 1 for SE events (indicating no evidence for exon skipping) and 0 for RI events (indicating no evidence for intron retention) to obtain the most accurate assessment of differential alternative splicing in P/LP carriers versus non-carriers. In cases where multiple exon skipping events were reported for the same exon but with different flanking exons, we retained the event with the greatest PSI difference in the P/LP carrier versus non-carriers. Significant germline P/LP variant-associated differential splicing events were defined as events with |PSI z-score| ≥ 2 and |PSI sample - mean PSI| > 0.05, where the P/LP variant was <250bp from the splice junction.

### DNA Methylation

Matched tumor Illumina 450K or EPIC 850K Infinium Human Methylation Bead Array data from the PBTA-CBTN Open Access CAVATICA project (https://cavatica.sbgenomics.com/u/cavatica/pbta-cbttc/files-premium?path=CBTN-Methylation) were processed to obtain beta values as previously described in the OpenPedCan project^26^. Briefly, IDATs were processed using the minfi Bioconductor package, funnorm normalization, and probes containing common SNPs in dbSNP (minor allele frequency > 1%) were removed. We calculated detection p-values using minfi and filtered out probes with signal that was indistinguishable from background (p<0.05). Separately as a part of OpenPedCan, the IDATs were run through the DKFZ brain classifier version 12.6^73^ and/or the NIH classifier v2 to obtain tumor classifications. To determine if P/LP carriers exhibited somatic differential methylation of affected CPG, we calculated beta value z-scores of each probe associated with that CPG in the same manner as described for gene expression. Significant differential methylation was defined as |sample beta value z-score| ≥ 2 and |sample beta value - mean beta value| > 0.05. We also calculated mean beta values across all measured probes for each patient as an estimate of relative global methylation across individuals.

### Proteomics

Proteomics data from the Clinical Proteomic Tumor Analysis Consortium (CPTAC PBTA, pediatric, Proteomic Data Commons PDC000180) and Project HOPE (adolescent and young adult HGG) were obtained from OpenPedCan release v15^26,74^. Sample-level protein abundance z-scores in P/LP carriers were calculated relative to other samples of the same histology were calculated as previously described. Significant differential protein abundance was defined as |sample protein abundance z-score| ≥ 2.

### Tumor mutational signatures analysis

We utilized mutational signatures generated from the v15 release of OpenPedCan^26^. Mutational signature analysis was performed using the deconstructSigs^75^ R package function `whichSignatures()`, input consensus SNV file, and COSMIC v3.3 mutational signatures database as described in the Open Pediatric Cancer Project^26^. Overall MMR deficiency exposure was calculated as the sum of the following SBS signatures: SBS6, SBS14, SBS15, SBS20, SBS21, SBS26, and SBS44. We performed Mann Whitney-U tests to assess differences in mutational signature exposure weights between samples with and without germline P/LP variants in DNA repair genes. Exposure weight differences were plotted using ggplot2 `geom_bar()` function^76^. Differences in mutational signatures exposure weights in >2 groups were assessed using one-way analysis of variance (ANOVA) and post-hoc pairwise comparisons, and were plotted using ggplot2 `geom_violin()` function^76^.

### Tumor Mutation Burden

We utilized tumor mutation burden (TMB) calculations from the v15 release of the Open Pediatric Cancer Project^26^. Briefly, coding-only TMB was computed using high- or moderate-impact nonsynonymous variants from consensus SNV/MNV calls, i.e. those detected by at least two of four variant callers or hotspot mutations from one variant caller. For WGS and WXS samples, only variants within coding regions defined by GENCODE v39 were counted, and TMB was calculated by dividing these counts by the size of the genome or exome coding regions surveyed by the variant callers.

### Survival analyses

We performed Kaplan-Meier analyses of overall and event-free survival (OS and EFS, respectively) to compare outcomes of patients with and without germline P/LP variants in CPGs. Patient events that were included in EFS calculations were as follows: tumor progression, recurrence, and second malignancy, and death due to disease. We generated Cox proportional-hazards regression models to identify variables that were predictive of outcome. Variables considered included presence of germline P/LP variant in a CPG, molecular subtype, extent of tumor resection, reported gender, and age at diagnosis. The cohort analyzed included all those for whom data were available for all variables in the model (n=654 for OS, n=652 for EFS). Survival analyses were run using the survival R package^77,78^.

## Inclusion and Ethics Statement

Human data analyzed for this manuscript was obtained through an approved data use agreement and utilized with patient consent for general research use.

## Data availability

All pediatric brain tumor raw data are available upon request from the database of Genotypes and Phenotypes (dbGaP), accession number phs002517.v4.p2. Proteomics raw data from the Clinical Proteomic Tumor Analysis Consortium (CPTAC PBTA, pediatric) are openly available from the Proteomic Data Commons PDC000180. Methylation array data are openly available from the PBTA-CBTN Open Access CAVATICA project (https://cavatica.sbgenomics.com/u/cavatica/pbta-cbttc/files-premium?path=CBTN-Methylation). All processed somatic data used in this study were derived from the OpenPedCan project^26^ v15 data release^79^, which can be found through https://github.com/d3b-center/OpenPedCan-analysis or CAVATICA https://cavatica.sbgenomics.com/u/cavatica/opentarget.

## Code availability

All original code and manuscript figures and tables are available at https://github.com/diskin-lab-chop/pbta-germline-somatic.

## Author Contributions

Conceptualization: SJD, SM, AJW, MB, KAC, RJC, JLR, RSK

Methodology: RJC, RSK, ZV, MAB, SP, JLR, SJD, ASN, SW, JMD, AS

Software: MAB, SP, JMD, AS

Validation: ASN, AC, RSK, RJC, MAB, ZG, ECCC, PJS, JLR, SJD

Formal Analysis: RJC, RSK, MAB, SP, BZ, CZ, JLR

Investigation: RJC, RSK, SJD, JLR, MB, SM, SWM, KAC, ML, SW, AJW

Resources: PBS, ACR, SJD, JLR, AJW

Data Curation: JLR, JLM, RJC, ZG, EB

Writing - Original Draft: RJC, JLR, SWM

Writing - Review & Editing: JLR, SJD, AJW, SW, SP, MB, SM, RJC, KAC, EG

Visualization: RSK, RJC, JLR, ASN

Supervision: JLR, SJD

Project Administration: JLR, SJD

Funding Acquisition: SJD, JLR, ACR

## Acknowledgements

The authors thank the patients and families for donating tissue to the pediatric oncology community and the Children’s Brain Tumor Network and Pacific Pediatric Neuro-Oncology Consortium for generating the sequencing data used for this work. This work was funded in part by National Institutes of Health (NIH) grants R03CA287169 (SJD, JLR), R03OD036498 (JLR), U24OD038422 (SJD, JLR), R03CA230366 (SJD), the NIH Kids First Cloud Credits Program (SJD, JLR), the Children’s Brain Tumor Network, the Chad Tough Foundation, and the anonymous private investors to the Children’s National Hospital Brain Tumor Institute.

