## Supplemental Information for "Germline pathogenic variation impacts somatic alterations and patient outcomes in pediatric CNS tumors"

**Supplementary Information**

[Supplementary Table 9. AutoGVP results of germline P/LP SNVs/indels of low allele frequency (0.08<VAF<0.2) (Excel file).	3](#_Toc203570215)

### I. Supplementary Tables

#### Supplementary Table 1. CNS tumor predisposition syndromes defined by WHO 2021 Classification (Excel file).

#### Supplementary Table 2. Pediatric CNS tumor patient clinical and demographic data (Excel file).

#### Supplementary Table 3. Cancer predisposition gene chromosomal locations, associated syndromes and cancer types, and modes of inheritance (Excel file).

Supplementary Table 4. Germline P/LP variant summary (Excel file). **A)** P/LP SNV/indel summary from AutoGVP. **B)** Germline variants upgraded to P/LP based on published functional evidence supporting loss of function. **C)** P/LP structural variant summary.

Supplementary Table 5. CPG P/LP variants identified P/LP carrier-enriched molecular subtypes (Excel file). **A)** Subependymal giant cell astrocytoma, **B)** SHH-activated medulloblastoma, **C)** H3-wildtype, TP53-altered HGG, and **D)** pineoblastoma.

Supplementary Table 6. Demographic and clinical features by CPG P/LP carrier status and histology (Excel file). **A)** Low-grade glioma, **B)** medulloblastoma, **C)** mixed glial-neuronal tumors, **D)** ependymoma, **E)** high-grade glioma, **F)** craniopharyngioma, **G)** AT/RT, **H)** meningioma, **I)** DIPG or DMG, **J)** mesenchymal tumors, **K)** neurofibroma plexiform, **L)** non-neoplastic tumor, **M)** germ cell tumor, **N)** schwannoma, **O)** choroid plexus tumor, **P)** other tumor cohorts.

Supplementary Table 7. Summary of predisposition syndrome-associated gene P/LP variants (Excel file). A) SNVs/indels and B) structural variants and concordance with clinical reports.

#### Supplementary Table 8. Germline variants of uncertain significance (VUS) and low variant allele frequency (VAF) P/LP variants in patients with clinically-reported syndromes but no associated P/LP variant (Excel file).

#### Supplementary Table 9. AutoGVP results of germline P/LP SNVs/indels of low allele frequency (0.08≤VAF<0.2) (Excel file).

Supplementary Table 10. P/LP variant burden testing in PBTA versus Penn Medicine BioBank (PMBB) and gnomAD cancer-free control cohorts (Excel file). Variant burden testing in **A)** all cancer predisposition genes (CPGs), **B)** each CPG, **C)** KEGG pathway gene sets, **D)** DNA repair pathway gene sets. Variant burden testing by histology in **E)** all CPGs, **F)** individual CPGs, **G)** DNA repair gene sets.

Supplementary Table 11. Summary of gene-level somatic alterations associated with germline P/LP variation (Excel file). **A)** germline P/LP SNVs/indels and **B)** germline structural variants in the same gene.

#### Supplementary Table 12. Summary of germline CPG P/LP variant-associated differentially methylated Infinium 850K EPIC array probes in matched tumors (Excel file).

#### Supplementary Table 13. Summary of germline CPG P/LP variant-proximal splice events in matched tumors (Excel file).

Supplementary Table 14. COSMIC single base substitution (SBS) mutational signatures exposures in HGG tumors of mismatch repair gene P/LP carriers versus non-carriers (Excel file). Statistics were derived from Wilcoxon rank sum tests.

#### Supplementary Table 15. Summary of tumor mutation – mean methylation beta value correlations by pediatric CNS tumor histology (Excel file).

Supplementary Table 16. Median event- free survival (EFS) and overall survival (OS) in germline CPG P/LP carriers versus non-carriers by tumor histology and molecular subtype (Excel file). Hazard ratio, p-values, and confidence intervals were derived from cox proportional hazards models.

### II. Supplementary Figures

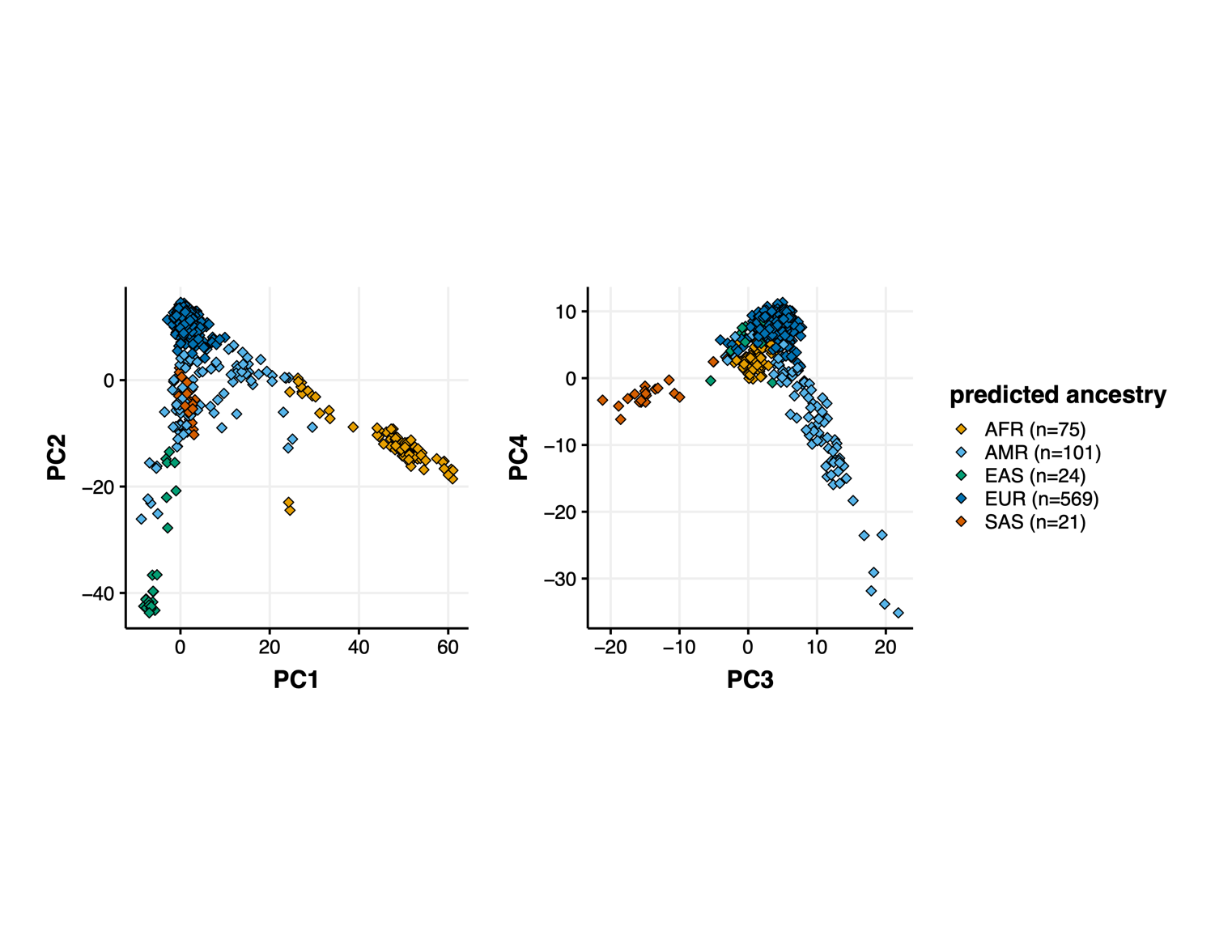

**Supplementary Figure 1. Genetic ancestry prediction in pediatric CNS tumor cohort.** Pediatric CNS tumor cohort principal components derived from Somalier genetic ancestry prediction. Samples are colored based on ancestry superpopulation assignment. AFR=African, AMR=admixed American, EAS=East Asian, EUR=European, SAS=South Asian.

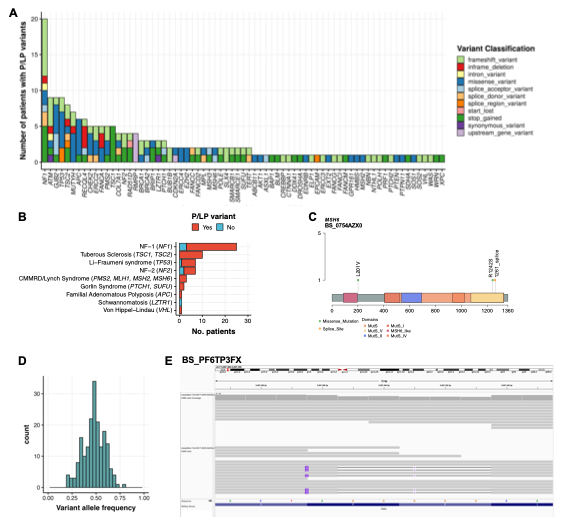

**Supplementary Figure 2. P/LP variant distribution in pediatric CNS tumor cohort. A.** Number of patients with identified germline CPG P/LP variants by gene and variant classification. **B.** Number of patients with cancer predisposition syndromes found to harbor P/LP variants in associated genes. **C.** A patient with pediatric H3 wildtype high-grade glioma and clinically-reported constitutional mismatch repair deficiency syndrome (CMMRD) harbors two missense and one splice site *MSH6* germline P/LP variants. **D.** Histogram of germline CPG P/LP variant allele frequencies. **E**. Integrated Genomics Viewer (IGV) display confirming the presence of two *PMS2* InDels on the same allele in a patient with medulloblastoma.

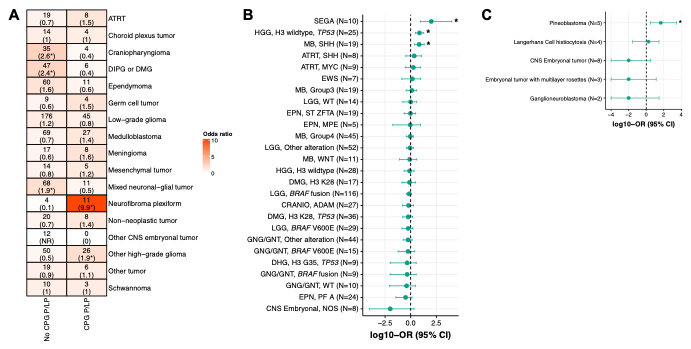

**Supplementary Figure 3. P/LP carrier distribution among CNS tumor histologies, molecular subtypes, and predisposition syndromes. A.** Heatmap displaying number of patients in each histology and CPG P/LP carrier status, with odds ratios in parentheses. Odds ratios for each histology were derived from contingency tables indicating P/LP carrier status (carrier or non-carrier) and membership in histology (yes or no). *FDR < 0.05. NR = not reportable. **B-C.** Dot plot of P/LP carrier enrichment odds ratios across **B)** CNS tumor molecular subtype cohort patients and **C)** tumor histologies classified into “Other tumor” histology group. *FDR < 0.05.

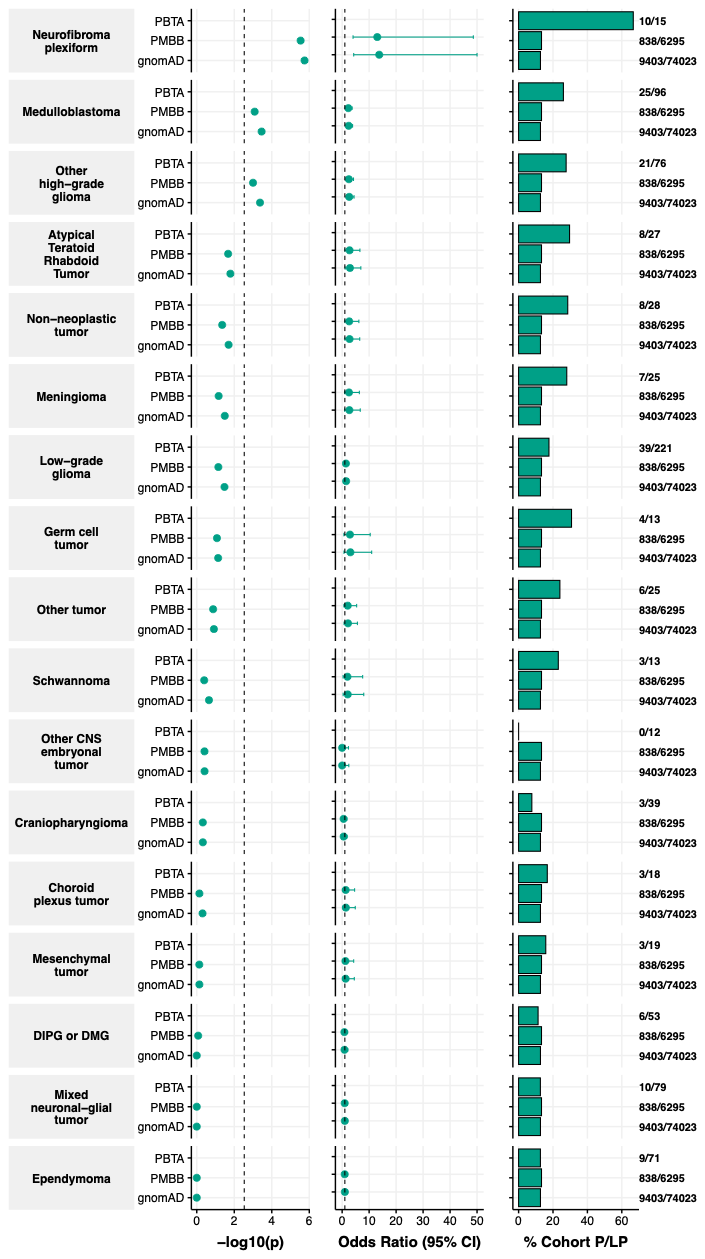

**Supplementary Figure 4. CPG P/LP variant burden among pediatric CNS tumor histologies.** Odds ratios and associated p-values of CPG P/LP variant burden among CNS tumor histology cohorts relative to PMBB and gnomAD cancer-free control cohorts. Dashed lines in p-value plots indicate Bonferroni-adjusted significance thresholds.

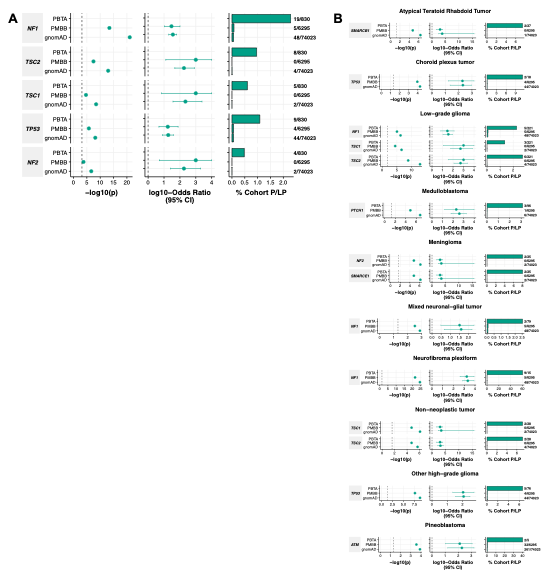

**Supplementary Figure 5. Gene-level P/LP variant burden in pediatric CNS tumor histologies. A-B.** Odds ratios and associated p-values of P/LP variant burden in significantly enriched cancer predisposition genes (CPGs) among **A)** full pediatric CNS tumor cohort and **B)** tumor histology cohorts relative to PMBB and gnomAD cancer-free control cohorts. Dashed lines in p-value plots indicate Bonferroni-adjusted significance thresholds.

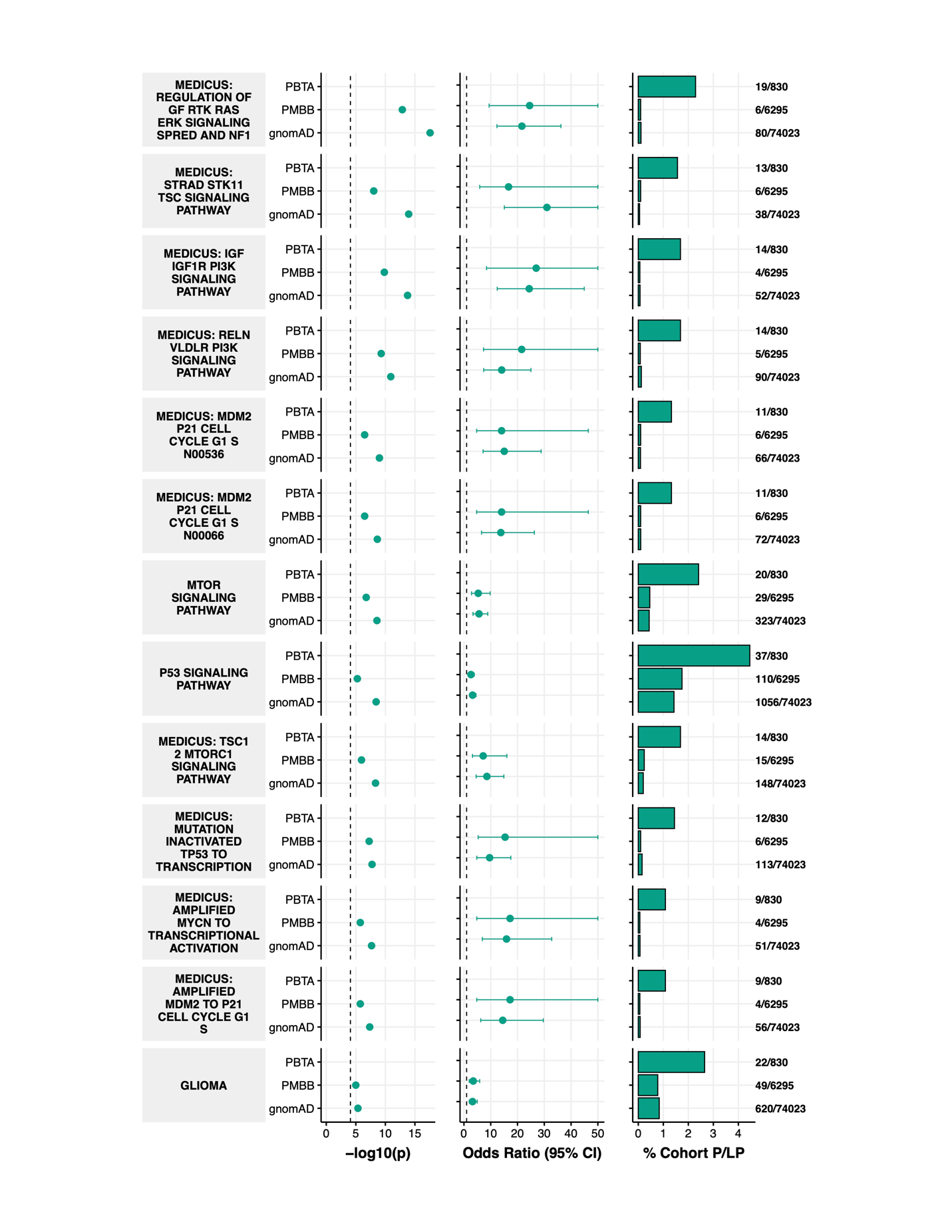

**Supplementary Figure 6. KEGG pathway-level P/LP variant burden in pediatric CNS tumor cohort.** Odds ratios and associated p-values of KEGG pathway gene P/LP variant burden in CNS tumor cohort relative to PMBB and gnomAD cancer-free control cohorts. Dashed lines in p-value plots indicate Bonferroni-adjusted significance thresholds.

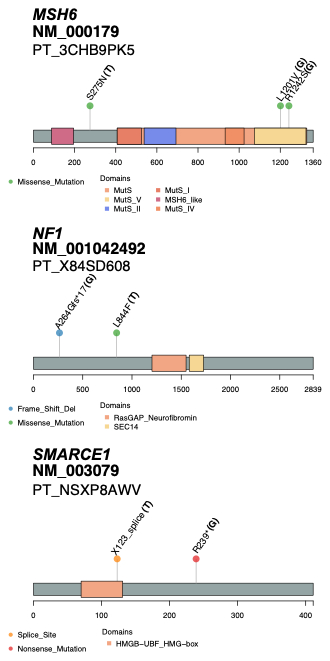

**Supplementary Figure 7. Putative deleterious SNV/indel second hits in germline CPG P/LP carriers.** Lollipop plots displaying germline P/LP variants (“G”) and somatic loss-of-function SNVs/indels (“T”) in matched tumors of *MSH6*, *NF1*, and *SMARCE1* P/LP carriers.

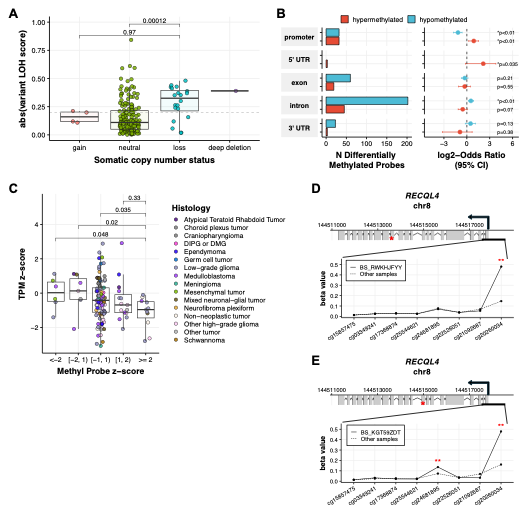

**Supplementary Figure 8. Somatic loss-of-heterozygosity and differential methylation in germline CPG P/LP carriers. A.** Somatic variant loss-of-heterozygosity (LOH) score (tumor VAF-germline VAF) by somatic copy number status. P-values are derived from Wilcoxon rank sum tests. **B.** Number of P/LP variant-associated differentially methylated probes by direction and gene feature, and associated Fisher’s exact test odds ratios representing extent of differential methylation enrichment in gene features. **C.** TPM z-score by z-score of most differentially methylated probe in promoter region. P-values are derived from Wilcoxon rank sum tests. **D-E.** Somatic *RECQL4* promoter-annotated probe beta values in two *RECQL4* P/LP carriers. Red asterisks in gene model plots indicate positions of P/LP variants, and asterisks in line plots indicate P/LP carrier z-score ≥ 2.

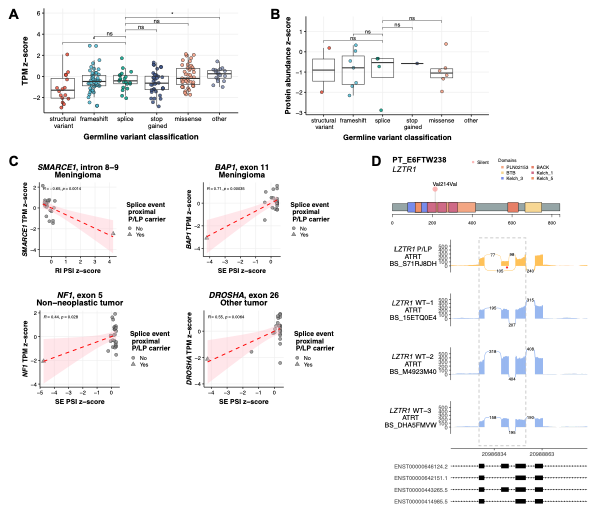

**Supplementary Figure 9. Germline P/LP variant-associated alternative splicing in pediatric CNS tumors. A-B.** Cancer predisposition gene (CPG) TPM z-scores **(A)** and protein abundance z-scores **(B)** in germline CPG P/LP carriers by germline variant classification. P-values are derived from Wilcoxon rank sum tests. **C.** Transcripts per million (TPM) z-scores against percent splice index (PSI) z-scores of splice events exhibiting significant differential alternative splicing in CPG P/LP carriers **D.** A *LZTR1* synonymous P/LP variant carrier with ATRT exhibits increased skipping of *LZTR1* exon 4 (chr22:20987503-20987583) relative to non-P/LP carriers with ATRT.

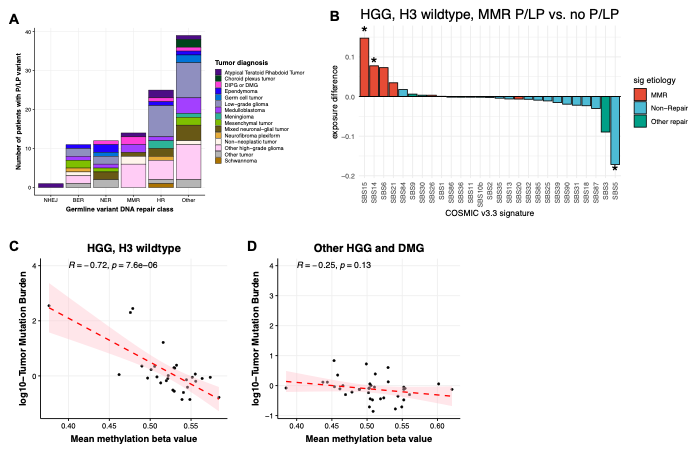

**Supplementary Figure 10. DNA repair gene P/LP variation in pediatric high-grade gliomas. A.** Number of patients with DNA repair gene P/LP variants by DNA repair pathway assignment and tumor histology. **B.** Bar plots indicating COSMICv3.3 mutational signature exposure weight differences in MMR gene P/LP carrier H3 wildtype HGG relative to non-P/LP carriers of the same tumor subtype. Asterisks indicate significant differences in signature exposure weights between groups (Wilcoxon rank sum p-value < 0.05). **C-D.** Scatter plots and Pearson correlation coefficients of tumor mutation burden versus sample mean methylation beta value in **C)** H3-wildtype HGG tumors and **D)** HGG tumors of other molecular subtypes.

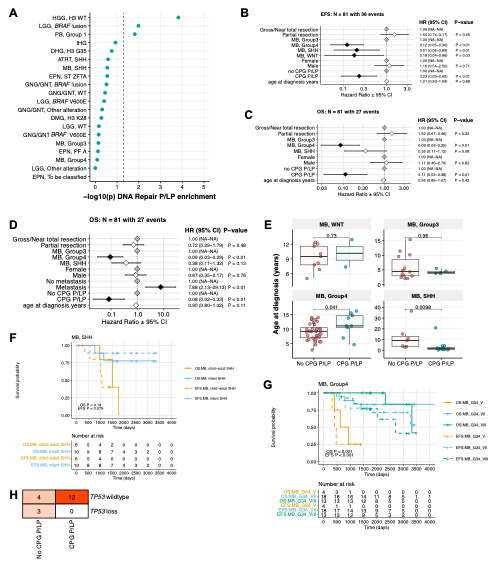

**Supplementary Figure 11. Molecular and clinical correlates with survival outcomes in germline CPG P/LP carriers. A.** DNA repair gene P/LP variant burden odds ratios in PBTA tumor molecular subtype cohorts relative to gnomAD cancer-free control cohort. **B-C.** Cox proportional hazards model forest plot of medulloblastoma event-free survival (EFS, **B**) and overall survival (OS, **C**) including extent of tumor resection, molecular subtype, CPG P/LP carrier status, and age at diagnosis covariates. **D.** Cox proportional hazards model forest plot of medulloblastoma OS including the same covariates as in **B** with the addition of metastasis status. **E.** Age at diagnosis by P/LP carrier status and MB molecular subtype. p-values are derived from Wilcoxon rank sum tests. **F.** Kaplan-Meier EFS and OS plot of SHH-activated MB cohort by SHH subtype (infant versus child-adult). **G.** Kaplan-Meier EFS and OS plot of Group 4 MB cohort by methylation subgroup. **H.** Distribution heatmap of somatic *TP53* alteration status by CPG P/LP carrier status in SHH-MB.

### III. Non-Author Collaborator Lists and Contributions

#### Penn Medicine BioBank

Penn Medicine BioBank, Department of Medicine, Perelman School of Medicine, University of

Pennsylvania, Philadelphia, PA, USA

**PMBB Leadership Team**

Daniel J. Rader, M.D., Marylyn D. Ritchie, Ph.D., Michael D. Feldman M.D.

Contribution: All authors contributed to securing funding, study design and oversight. All authors reviewed the final version of the manuscript.

**Patient Recruitment and Regulatory Oversight**

JoEllen Weaver, Afiya Poindexter, Ashlei Brock, Khadijah Hu-Sain, Yi-An Ko

Contributions: JW manage patient recruitment and regulatory oversight of study. AP, AB, KH, YK recruitment and enrollment of study participants.

**Lab Operations**

JoEllen Weaver, Meghan Livingstone, Fred Vadivieso, Ashley Kloter, Stephanie DerOhannessian, Teo Tran, Linda Morrel, Ned Haubein, Joseph Dunn

Contribution: JW, ML, FV, SD oversight of lab operations. ML, FV, AK, SD, TT, LM perform sample processing. NH, JD are responsible for sample tracking and the laboratory information management system.

**Clinical Informatics**

Anurag Verma, Ph.D., Colleen Morse, M.S., Marjorie Risman, M.S., Renae Judy, B.S.

Contribution: All authors contributed to the development and validation of clinical phenotypes used to identify study subjects and (when applicable) controls.

**Genome Informatics**

Anurag Verma Ph.D., Shefali S. Verma, Ph.D., Yuki Bradford, M.S., Scott Dudek, M.S., Theodore Drivas, M.D., PH.D.

Contribution: A.V., S.S.V. are responsible for the analysis design and infrastructure needed quality control genotype and exome data. Y.B. performed the analysis. T.D. and A.V. provide variant and gene annotations and their functional interpretation of variants.

#### Regeneron Genetics Center

Regeneron Genetics Center, Regeneron Pharmaceuticals Inc., Tarrytown, NY, USA

**RGC Management and Leadership Team**

Goncalo Abecasis, Aris Baras, Michael Cantor, Giovanni Coppola, Andrew Deubler, Aris Economides, Katia Karalis, Luca A. Lotta, John D. Overton, Jeffrey G. Reid, Katherine Siminovitch & Alan Shuldiner

**Sequencing and Lab Operations**

Christina Beechert, Caitlin Forsythe, Erin D. Fuller, Zhenhua Gu, Michael Lattari, Alexander Lopez, John D. Overton, Maria Sotiopoulos Padilla, Manasi Pradhan, Kia Manoochehri, Thomas D. Schleicher, Louis Widom, Sarah E. Wolf & Ricardo H. Ulloa

**Clinical Informatics**

Amelia Averitt, Nilanjana Banerjee, Michael Cantor, Dadong Li, Sameer Malhotra, Deepika Sharma & Jeffrey C. Staples

**Genome Informatics**

Xiaodong Bai, Suganthi Balasubramanian, Suying Bao, Boris Boutkov, Siying Chen, Gisu Eom, Lukas Habegger, Alicia Hawes, Shareef Khalid, Olga Krasheninina, Rouel Lanche, Adam J. Mansfield, Evan K. Maxwell, George Mitra, Mona Nafde, Sean O’Keeffe, Max Orelus, Razvan Panea, Tommy Polanco, Ayesha Rasool, Jeffrey G. Reid, William Salerno, Jeffrey C. Staples, Kathie Sun & Jiwen Xin

**Analytical Genomics and Data Science**

Goncalo Abecasis, Joshua Backman, Amy Damas, Lee Dobbyn, Manuel Allen Revez Ferreira, Arkopravo Ghosh, Christopher Gillies, Lauren Gurski, Eric Jorgenson, Hyun Min Kang, Michael Kessler, Jack Kosmicki, Alexander Li, Nan Lin, Daren Liu, Adam Locke, Jonathan Marchini, Anthony Marcketta, Joelle Mbatchou, Arden Moscati, Charles Paulding, Carlo Sidore, Eli Stahl, Kyoko Watanabe, Bin Ye, Blair Zhang & Andrey Ziyatdinov

**Research Program Management & Strategic Initiatives**

Marcus B. Jones, Jason Mighty & Lyndon J. Mitnaul
